# A systematic review of ambient heat and sleep in a warming climate

**DOI:** 10.1101/2023.03.28.23287841

**Authors:** Guillaume Chevance, Kelton Minor, Constanza Vielma, Emmanuel Campi, Cristina O’Callaghan-Gordo, Xavier Basagaña, Joan Ballester, Paquito Bernard

## Abstract

**Background:** Earlier reviews documented the effects of a broad range of climate change outcomes on sleep but have not yet evaluated the effect of ambient temperature. This systematic review aims to identify and summarize the literature on ambient temperature and sleep outcomes in a warming world.

**Methods:** For this systematic review, we searched online databases (PubMed, Scopus, JSTOR, GreenFILE, GeoRef and PsycARTICLES) together with relevant journals for studies published before February 2023. We included articles reporting associations between objective indicators of ambient temperature and valid sleep outcomes measured in real-life environments. We included studies conducted among adults, adolescents, and children. A narrative synthesis of the literature was then performed.

**Findings:** The present systematic review shows that higher outdoor or indoor ambient temperatures, expressed either as daily mean or night-time temperature, are negatively associated with sleep quality and quantity worldwide. The negative effect of higher ambient temperatures on sleep is stronger in the warmest months of the year, among vulnerable populations and in the warmest areas of the world. This result appears consistent across several sleep indicators and measures.

**Interpretation:** Although this work identified several methodological limitations of the extant literature, a strong body of evidence from both this systematic review and previous experimental studies converge on the negative impact of elevated temperatures on sleep quality and quantity. In absence of solid evidence on fast adaptation to the effects of heat on sleep, rising temperatures induced by climate change pose a planetary threat to human sleep and therefore human health, performance and wellbeing.

## Introduction

There is accumulating evidence that climate change is increasing health risks and notably heat-related illnesses and mortality.^1–5^ Beyond mortality, hotter ambient temperatures and heat extremes are associated with increased injuries,^6,7^ hospitalizations,^8^ mental health issues,^9^ health-care costs,^10^ as well as worsened cognitive performance,^11^ sentiment,^4^ labor productivity,^12^ and activity days.^13^ One proposed pathway of the association between ambient temperature and health outcomes is disrupted sleep.^14–16^ Shorter sleep duration, poor sleep quality and sleep disorders (e.g., insomnia, sleep apnea) are prospectively associated with the development of cardiovascular^17^ and metabolic diseases,^18^ cancer risks,^19^ mental health disorders,^20^ and accidents.^21^ Although the environmental causes of sleep disruption are multi-factorial (e.g., light and noise pollution),^22,23^ it is likely that rising ambient temperatures due to ongoing climate change will impair sleep in the hottest seasons of the year at a global scale, barring further adaptation.^14,24^

Exposure to hot and cold ambient thermal conditions demand the human body to mount a thermoregulatory response to maintain a core body temperature rhythm within the normal range required to support physiological functioning and sound sleep.^25–27^ Extreme heat can alter human core body temperature outside its normal range when air temperatures exceed that of fully vasodilated skin (35°C), with elevated heat-health risks apparent well-below this threshold.^3,28–30^ Further, sleep onset is closely coupled with night-time core body temperature decline.^25^ In hot sleeping environments, heat production can exceed heat loss beyond tolerable levels, increasing core body temperature and disturbing the natural sleep–wake cycle with increased wakefulness.^26^ In daily life, ambient heat can impact core body temperature (and thus sleep) through at least two plausibly interacting pathways: (*i*) direct exposure during the day imposing thermal strain, cardiovascular strain and/or dehydration which may carry over into the nocturnal resting period,^3^ and (*ii*) exposure at night via a combination of nighttime ambient weather conditions and environmental heat transfer (i.e., the energy accumulated in the built environment, conducted and re-emitted in the bedroom at night) reducing the thermal gradient between the body and ambient environment.^26,31,32^

The largest investigation of the effect of ambient temperature on sleep thus far – a study based on billions of repeated sleep measurements from sleep-tracking wristbands collected in 68 countries over two years – found that increased nighttime ambient temperature shortens sleep duration, primarily through delayed sleep onset, with stronger negative effects during summer months, in lower-income countries, in warmer climate regions, among older adults, females and after controlling for individual and spatiotemporal confounders.^24^ These results confirmed those from two previous large-scale national analyses of self-reported sleep outcomes from the United States,^33,34^ the one including the most representative sample showing that a +1°C increase in monthly nighttime ambient temperatures produces an increase of approximately three nights of subjective insufficient sleep per 100 individuals per month.^33^ Minor et al. (2022) and Obradovich et al. (2017) also included climate change impact projections, and both estimated that rising ambient temperatures may negatively impact human sleep through the year 2100 under both moderate and high greenhouse gas concentration scenarios, with impacts scaling with the level of emissions barring further adaptation.^24,33^

Critically, climate change and other anthropogenic environmental changes related to increased heat exposure, such as urban heat island, are altering outdoor ambient temperatures where populations reside. Although sound and sufficient slumber underpins human functioning, the current prevalence of insufficient or poor sleep is already elevated in high-income countries (e.g., above 50% in the US, 31% in Europe and 23% in Japan).^35^ Although insufficient sleep prevalence estimates are relatively lower for low (7%)- and middle (8.2%)-income countries,^36^ a recent study found that habitants from those countries disproportionally suffered greater sleep loss due to heat,^24^ suggesting heightened vulnerability to elevated temperatures (i.e., reduced or missing access to personal or collective cooling strategies).^37^ In parallel, the 1.5°C global temperature threshold above the pre-industrial climate is expected to be exceeded by 2040 under most scenarios of the Intergovernmental Panel for Climate Change, including the increasingly plausible “Shared Socioeconomic Pathway” SSP2-4.5.^38^ Although there is some evidence of adaptation to increasing temperatures in high-income countries for heat-attributed mortality,^39^ temperature projections for the coming decades remain concerning for both current and future generations.^40–44^ In this context, having a precise and comprehensive understanding of the influence of ambient temperature and extreme heat on sleep is crucial.

Earlier reviews documented the effects of a broad range of climate change outcomes on sleep (see^14^ which included six studies about the specific role of ambient temperature and heat), the influence of the bed micro-environment on sleep physiology,^26,45,46^ as well as the specific role of humidity in sleep regulation measured mostly in laboratory settings.^47^ However, no previous systematic review has described the state of the literature on the effect of ambient temperature on sleep in humans under real-life conditions (i.e., observational studies conducted in real-life environments), in contrast with laboratory studies that experimentally manipulate both behavior and temperature in the micro-environment.^48–50^ This systematic review aims to identify and summarize the literature on ambient temperature, notably heat, and sleep outcomes in a warming world. Specifically, we aim to synthesize the available evidence and research gaps on this topic to inform researchers seeking to explore new facets of the temperature-sleep association, as well as decision makers and interventionists trying to promote climate adaptation.

## Methods

Methods for collecting and summarizing data met the standards of the Preferred Reporting Items for Systematic Reviews and Meta-Analyses (PRISMA) guidelines.^51^ The study protocol was registered in PROSPERO (CRD42021284139).

### Inclusion and Exclusion Criteria

Studies were included if they: (*i*) reported associations between objective indicators of ambient temperature and valid sleep outcomes measured in real-life, environments; (*ii*) included adults, adolescents, or children; and (*iii*) were peer-reviewed. Eligible measures of sleep included: self-reported (subjective) sleep questionnaires; accelerometer-based actigraphy and commercial-grade activity monitors (e.g., fitness bands and sleep-tracking wearable devices); sleep sensors and polysomnography. This selection criteria thus excluded sleep-adjacent articles such as those interested in the association between ambient temperature and the prescription of hypnotics, which are only indirectly related to sleep issues.^52^ Eligible measures for temperature were narrowed to objective records via weather stations, climate reanalysis data or indoor temperature sensors; this criterion excluded articles interested in the associations between sleep outcomes and seasons without measuring ambient temperature,^53^ subjective perceptions of temperature^54^ or using climate classifications (i.e., Koppen’s weather climate classification).^55^ We excluded articles with valid measures of both sleep and temperature when the association between the two outcomes was insufficiently reported (e.g., association plotted with insufficient details to be numerically interpreted and reported),^56^ or simply not tested (i.e., some articles include measures of temperature and sleep but focus on other outcomes).^57,58^ Because we focused on ambient temperature and sleep outcomes measured in real-life contexts, experimental studies manipulating in-laboratory temperatures were also excluded.^48–50^ Beyond heat manipulation, those experimental studies often include invasive skin and rectal temperature measures as well as behavioral constraints and thus can neither be considered ecologically valid in the context of daily life nor the present review.

### Data Sources and Searches

Studies were identified by searching PubMed, Scopus, JSTOR, GreenFILE, GeoRef and PsycARTICLES between March and April 2022, and then updated in February 2023. Search strategies and algorithms are available in the supplemental material. Relevant reviews and articles cited in the introduction were also scanned. Specific journals were also inspected, in the sleep literature (e.g., *Sleep; Sleep Medicine*, *Journal of Sleep Research*) and the environmental health domain (e.g., *Environment International*, *Environmental Research Letters*, *Environmental Health Perspectives, The Lancet Planetary Health*). After duplicates were removed, titles and abstracts of all studies identified were examined independently by three authors (GC, KM, PB) to determine those meeting the selection criteria.

### Data Extraction and Synthesis

An a priori data extraction form was developed and tested with 5 articles. Data were coded from each paper by three coders (KM, EC, PB) and double checked by a single coder (GC). The following information was extracted: first author’s name, sample region and period, study design, main research question, estimation strategy, temperature and sleep assessment methods and outcomes, the inclusion of relevant control variables, main results, conclusion, and relevant additional information. A narrative synthesis of the literature was then performed. We did not perform meta-analyses given the diversity of study designs, temperature and sleep outcomes and the statistical strategies used. A preliminary synthesis was conducted by the first authors (GC, KM) and then discussed and revised by all authors.

### Quality rating and risk of bias

We used the items from the Quality Assessment Tool for Observational Cohort and Cross-Sectional Studies^59^ as a basis to develop a custom list of 14 quality criteria relevant to the question of ambient temperature and sleep. For each criterion, we indicated whether or not the study met the requirement (i.e., binary outcome, yes/no). Criteria are displayed in Table 1 below.

**Table 1.** Quality criteria

|  |  |
| --- | --- |
| Item 1 | For exposures that can vary in amount or level, did the study examine different levels of the exposure as related to the outcome and allow for a non-linear or semi-parametric response? |
| Item 2 | Was the association between temperature and sleep assessed more than once over time at the within-participant level? |
| Item 3 | Was within-participant variance accounted for in the analyses? |
| Item 4 | Were sleep and temperature outcomes measured over the whole year? |
| Item 5 | Were seasons and/or day length measured and adjusted statistically for their impact on the relationship between exposure(s) and outcome(s)? |
| Item 6 | Were temporal variables, such as the day of the study, day of the year or day of the week, measured and adjusted statistically for their impact on the relationship between exposure(s) and outcome(s)? |
| Item 7 | Was humidity measured and adjusted statistically for its impact on the relationship between exposure(s) and outcome(s)? |
| Item 8 | Was wind speed measured and adjusted statistically for its impact on the relationship between exposure(s) and outcome(s)? |
| Item 9 | Was cloud cover measured and adjusted statistically for its impact on the relationship between exposure(s) and outcome(s)? |
| Item 10 | Was precipitation measured and adjusted statistically for its impact on the relationship between exposure(s) and outcome(s)? |
| Item 11 | Were indoor and outdoor temperatures measured and analyzed together? |
| Item 12 | Was the association between the exposure and the outcome tested at different lags? |
| Item 13 | Was the utilization of air conditioning, fans and/or other indoor personal cooling technologies measured and analyzed? |
| Item 14 | Was sleep measured with both subjective and device-derived indicators? |

## Results

### Descriptive findings

As depicted in the study flowchart (Figure 1), a first iteration resulted in a total of 1735 independent records. After screening title and abstract, 58 articles were inspected in closer detail and 27 articles were included in the present review.

**Figure 1.**
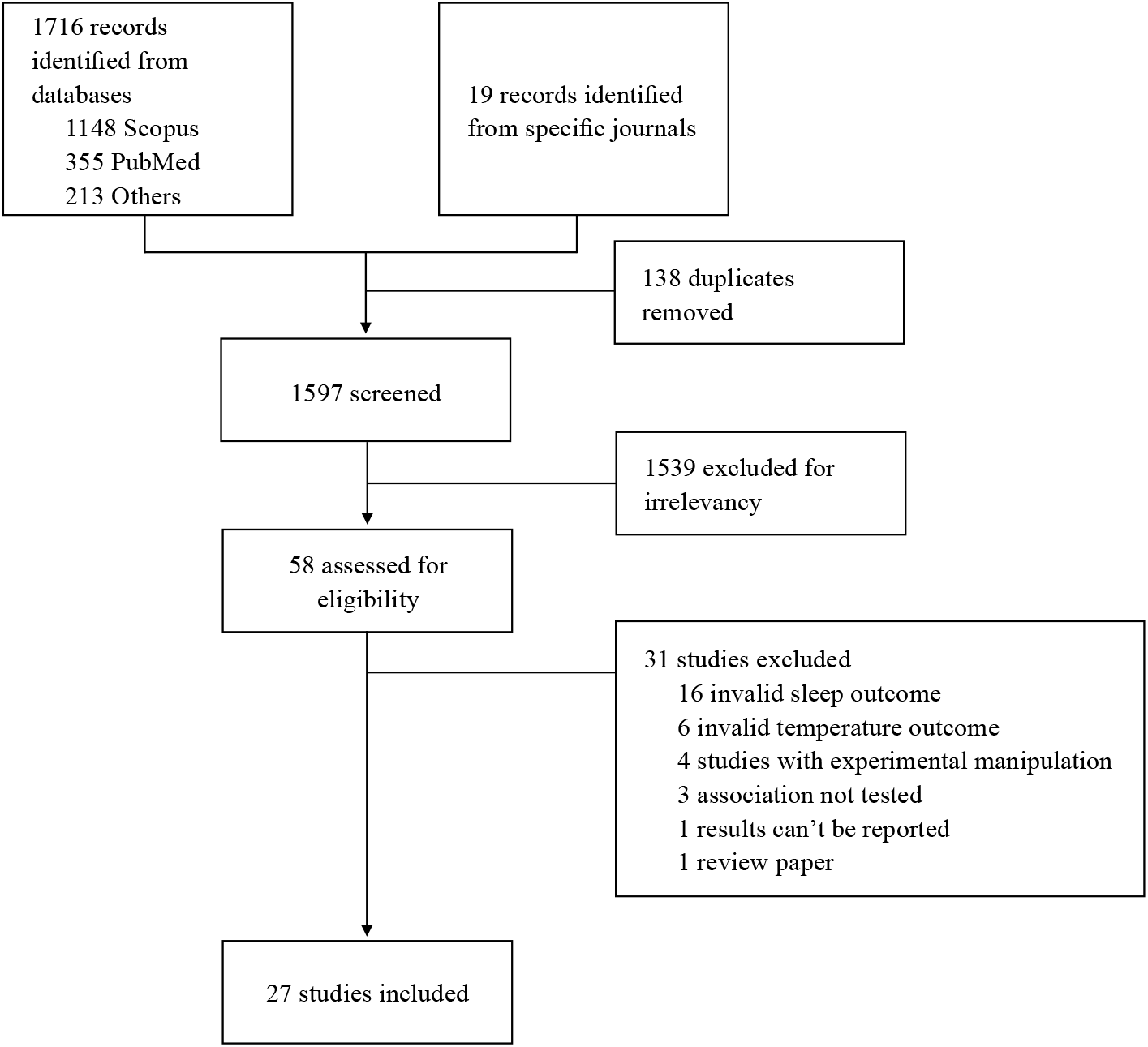
Study flowchart

Included articles were published in journals from diverse academic disciplines, including public health, sleep, physiology, engineering, economics, environmental science and medicine. The oldest article retrieved was published in 1992 and the most recent one 30 years later, in 2022. Figure 2 illustrates the countries in which the studies were performed; the US is the most represented country (42%). In terms of participants’ characteristics, studies included samples of various ages from ∼13 years old to nearly 80 years old. As for identified sex, most studies reported at least ∼30% female sample composition, while females were not included in two studies.^60,61^ All studies were correlational but six articles used intensive repeated measurements and appropriate statistical methods to estimate covariation between changes in temperatures and sleep outcomes within-participants, and thus are labelled as quasi-experimental studies here^24,60,62–65^.

**Figure 2.**
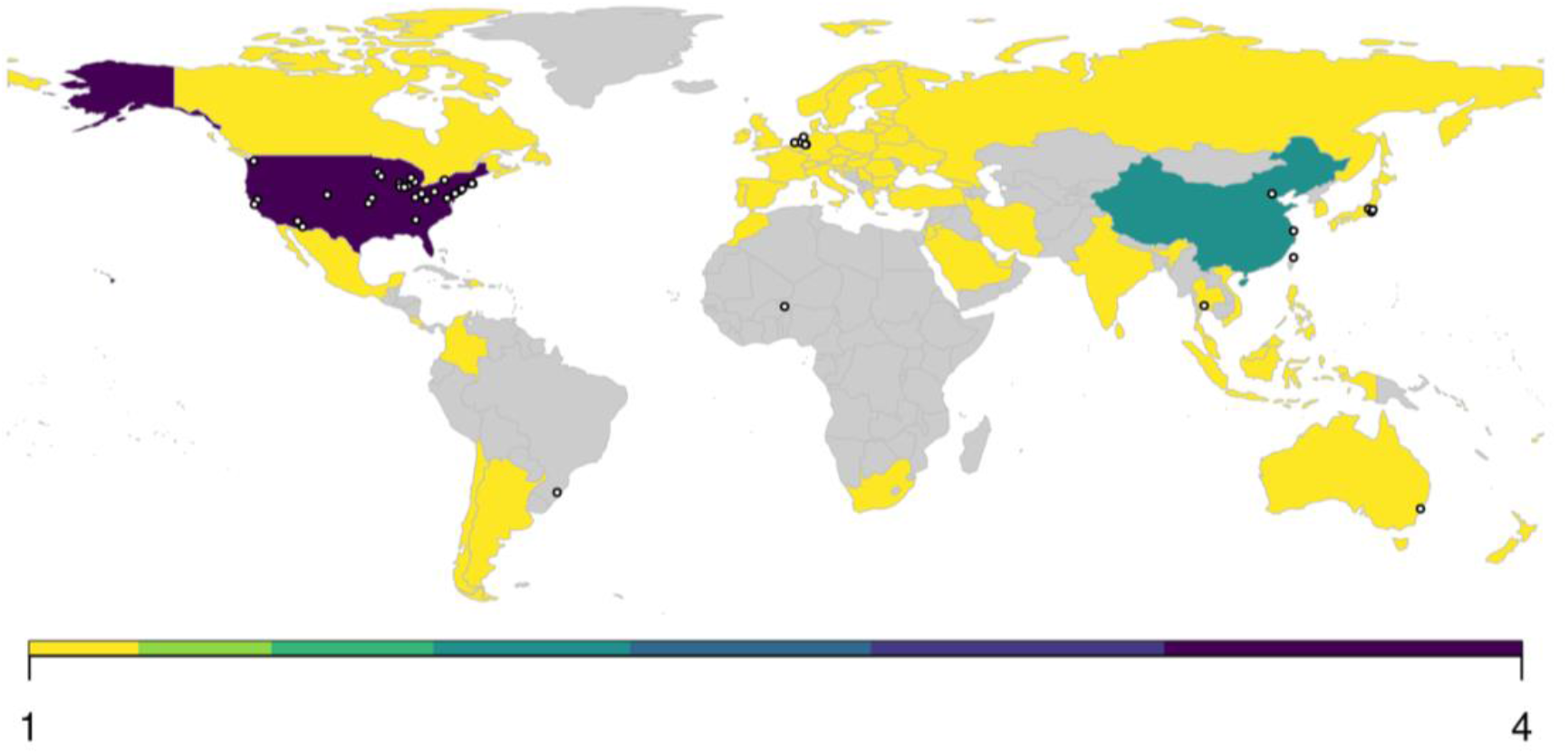
World map of included studies

Table 2 and Table 3 provide, respectively, a brief and detailed description (constrained by the scope of the current review) of each individual study included. Outdoor ambient temperature was measured via weather stations in 17 articles and indoor temperature was measured using local sensors installed in the home environment (e.g., HOBO temperature data logger, Thermochrons iButtons logger) in 11 articles. One study measured and reported results for both indoor (via local sensor) and outdoor (via weather station) temperature outcomes (see Table 2).^66^ Sleep measures showed a greater diversity of assessment methods with ten articles using self-reported questionnaires or diaries,^33,34,64–71^ eight studies using commercial activity monitors,^24,62,63,71–75^ five studies using polysomnography^61,69,76–78^, four studies using research-grade accelerometers^64,79–81^ and three studies using specific sleep sensors^60,82,83^. Beyond assessment methods, 15 articles used daily (24-hour) aggregated measures of temperature, the remaining 12 articles focused on average nighttime temperature (see Table 3, column “sample period”). For sleep, outcomes ranged from subjective sleep duration^34^, perception of insufficient sleep^33^ or sleep quality^70^ to accelerometer-derived signals providing information about sleep efficiency or wakefulness after sleep onset^64^ as well as sleep-related outcomes measured via polysomnography such as sleep stages^78^ and sleep disorders (e.g., episodes of sleep apnea).^76^

**Table 2.** Short summary of included studies

| Study | Country | Temperature assessment | Sleep assessment | Sleep outcome | Impact of increased temperature |
| --- | --- | --- | --- | --- | --- |
| An, 2018<br>N = 12,000 | China | Weather station | Questionnaire | Sleep duration | Positive |
| Cassol, 2012<br>N = 7,523 | Brazil | Weather station | PSG | AHI | Positive |
| Cedeño L., 2018<br>N = 44 | US | Local sensor | Activity monitor | Sleep duration | Negative |
| Cepeda, 2018<br>N = 1,166 | The Netherlands | Weather station | Research-grade accelerometer | Sleep duration | Negative |
| Hashizaki, 2018<br>N = 1,856 | Japan | Weather station | Other sensor | Sleep timing; WASO; SE | Negative |
| Lappharat, 2018<br>N = 68 | Thailand | Local sensor | Questionnaire + PSG | Subjective sleep; Sleep latency; Sleep duration; AHI | Negative |
| Li, 2020<br>N = 98 | US | Weather station | Research-grade accelerometer + Questionnaire | WASO; SE; Sleep duration; Subjective sleep; Sleep latency | Negative |
| Liu, 2022<br>N = 5,204 | Taiwan | Weather station | PSG | WASO; SE; AHI; sleep stages | Negative |
| Mattingly, 2021<br>N = 216 | US | Weather station | Activity monitor | Sleep duration and timing | Not explicit |
| Milando, 2022<br>N = 22 | US | Local sensor | Activity monitor | Sleep duration | Not significant |
| Minor, 2022<br>N = 47,628 | 68 countries | Weather station | Activity monitor | Sleep duration and timing | Negative |
| Montmayeur, 1992<br>N = 6 | Niger | Weather station | PSG | Sleep duration; SE; Sleep stages and awakenings | Not explicit |
| Mullins, 2019<br>N = 4,120,514 | US | Weather station | Questionnaire | Subjective sleep; Sleep duration | Negative |
| Obradovich, 2017<br>N = 765,000 | US | Weather station | Questionnaire | Subjective sleep | Negative |
| Ohnaka, 1995<br>N = 40 | Japan | Local sensor | Other sensor | Body movements | Negative |
| Okamoto, 2010<br>N = 19 | Japan | Local sensor | Research-grade accelerometer | Sleep timing; Sleep duration; WASO; SE | Negative |
| Pandey, 2005<br>N = 43 | US | Weather station | Questionnaire | Sleep latency; Awakenings; WASO; sleep duration | Negative |
| Quante, 2017<br>N = 669 | US | Weather station | Research-grade accelerometer | Sleep duration and timing; WASO; SE | Negative |
| Quinn, 2016<br>N = 40 | US | Weather station<br>+ Local sensor | Questionnaire | Subjective sleep | Negative |
| van Loenhout, 2016<br>N = 113 | The Netherlands | Local sensor | Questionnaire | Subjective sleep | Negative |
| Wang, 2022<br>N = 40,470 | China | Weather station | Questionnaire | Subjective sleep | Negative |
| Weinreich, 2015<br>N = 1,773 | Germany | Weather station | Other sensor | AHI | Negative |
| Williams, 2019<br>N = 51 | US | Local sensor | Activity monitor | Body movements | Negative |
| Xiong, 2020<br>N = 48 | Australia | Local sensor | Activity monitor | SE; Sleep stages | Negative |
| Xu, 2021<br>N = 41 | China | Local sensor | Activity monitor | SWS | Negative |
| Yan, 2022<br>N = 40 | China | Local sensor | Activity monitor;<br>Questionnaire | Sleep duration; REM; WASO;<br>SE; Light sleep; Deep sleep;<br>Subjective sleep | Negative |
| Zanobetti, 2009<br>N = 3,030 | US | Weather station | PSG | RDI | Negative |

**Abbreviations.** PSG: polysomnography; AHI: apnea-hypopnea index; WASO: wake after sleep onset; SE: sleep efficiency; TST: total sleep time; NREM: non-rapid eye movement; RDI: respiratory disturbance index.

**Table 3.**
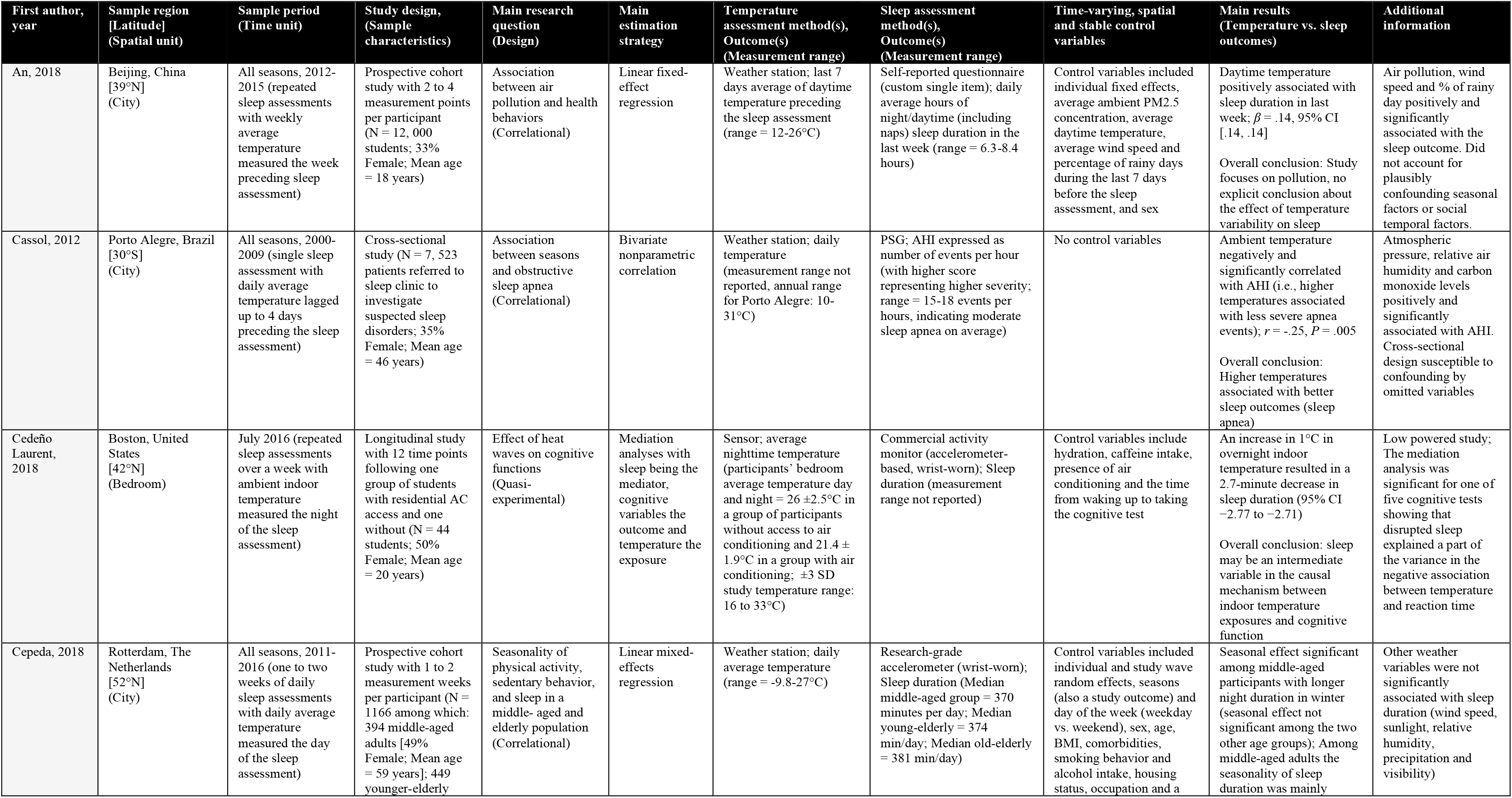
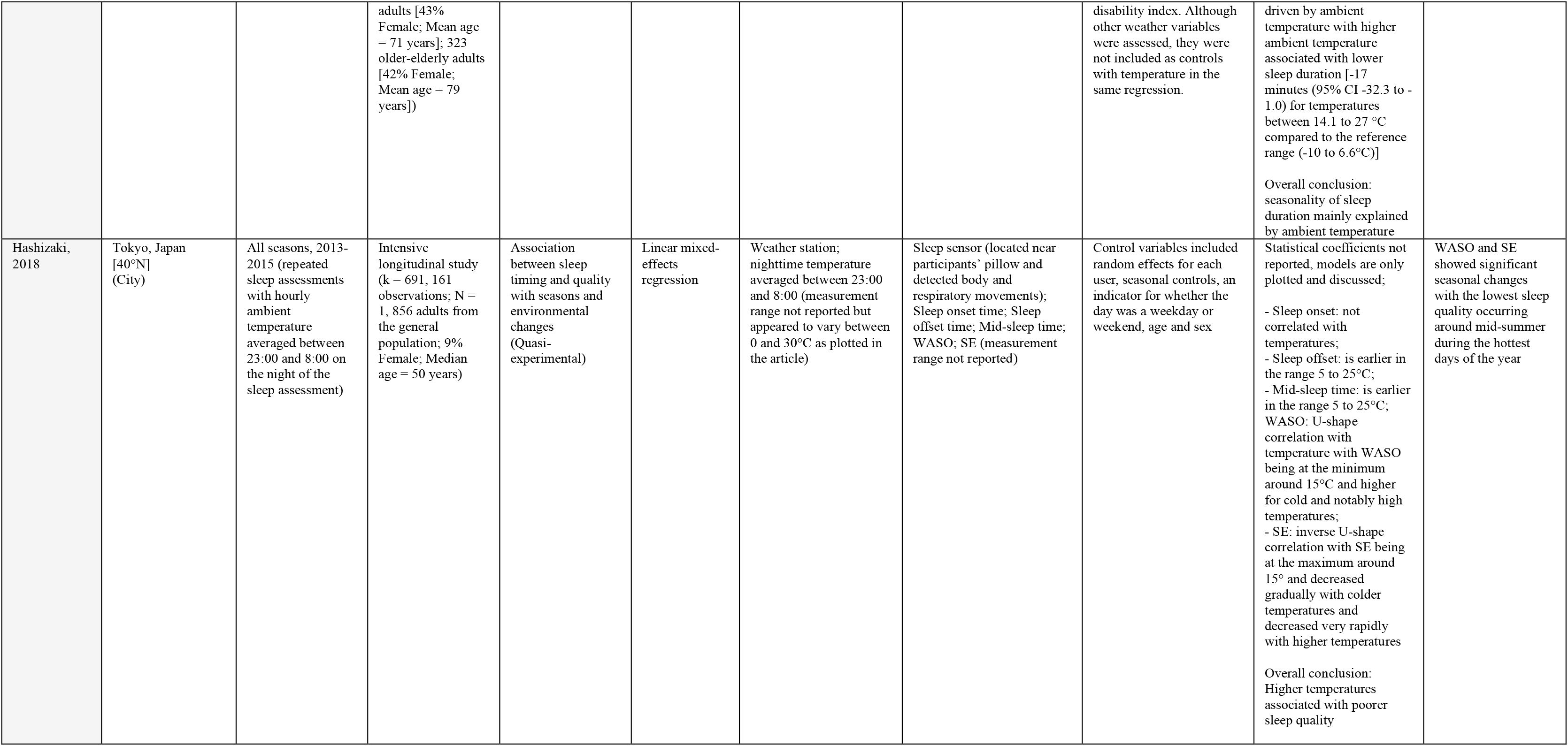
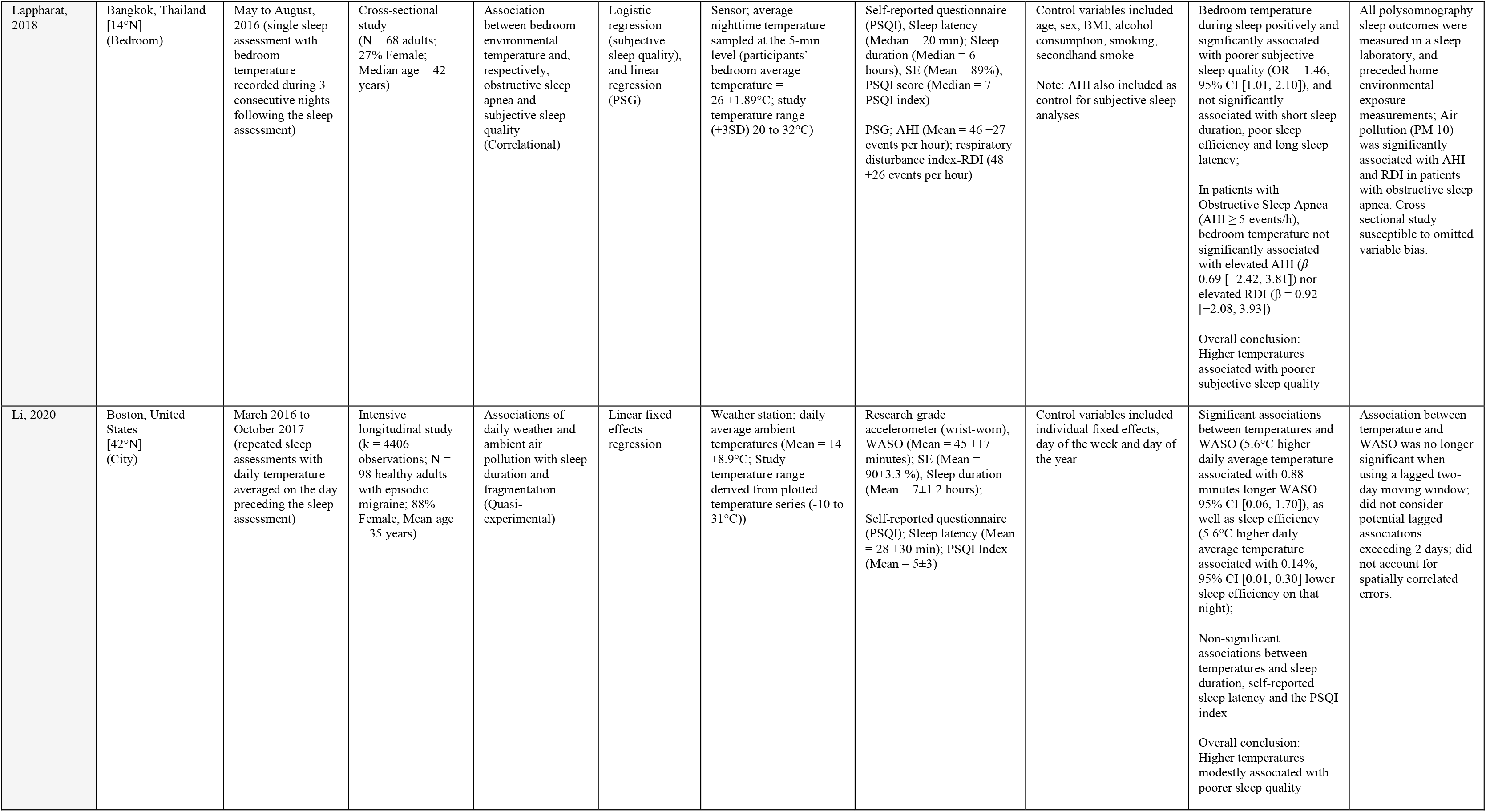
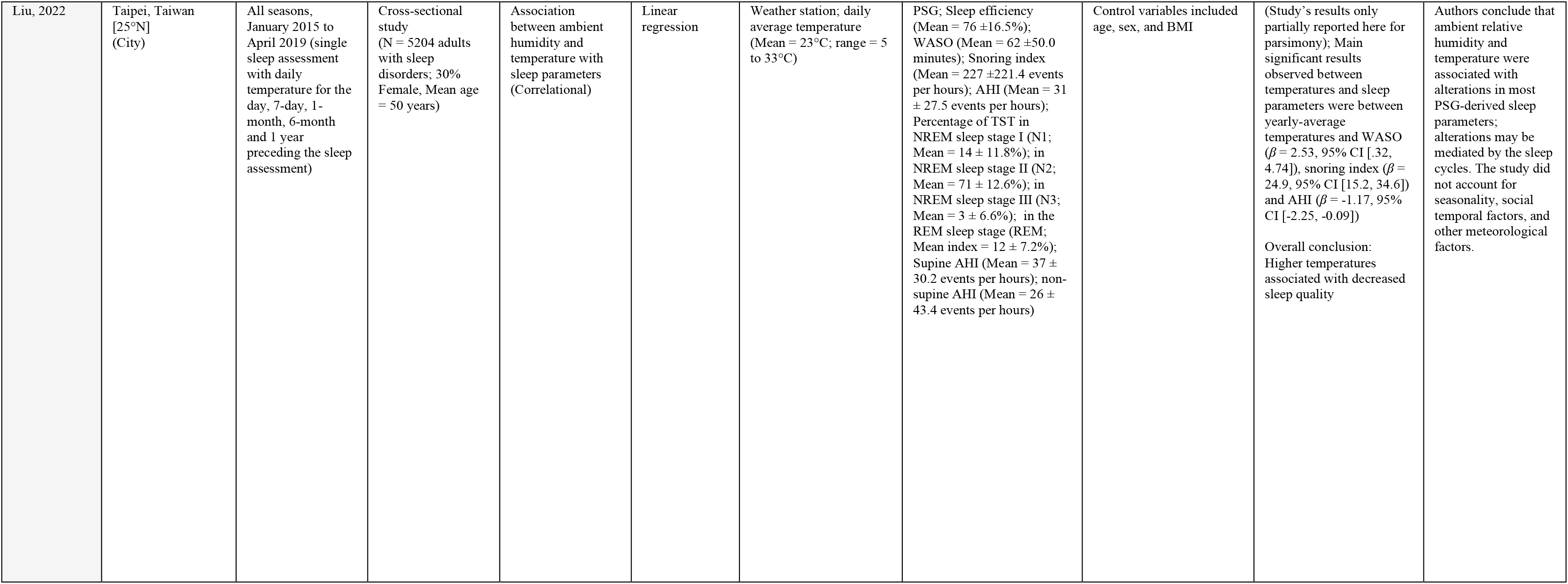
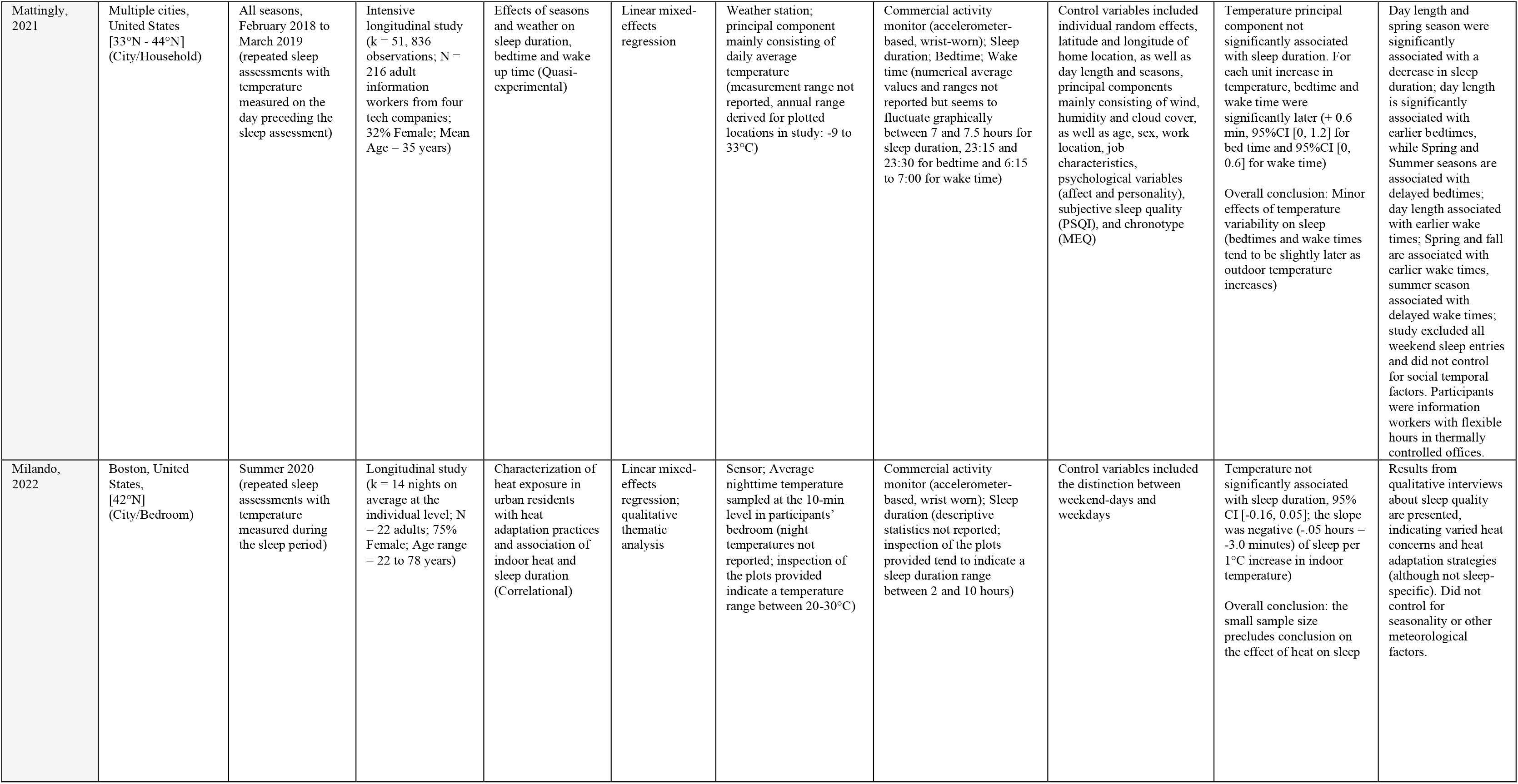
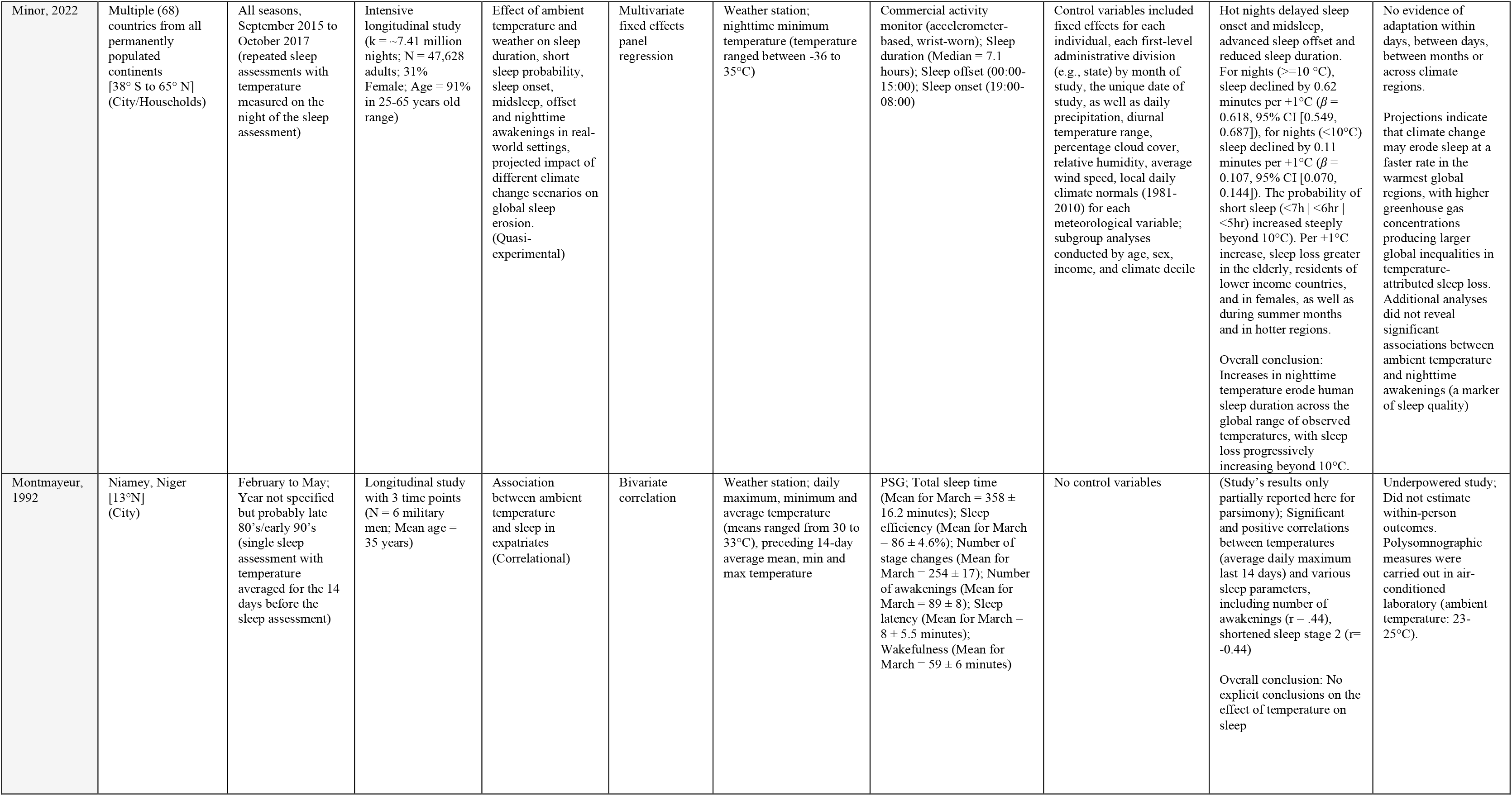
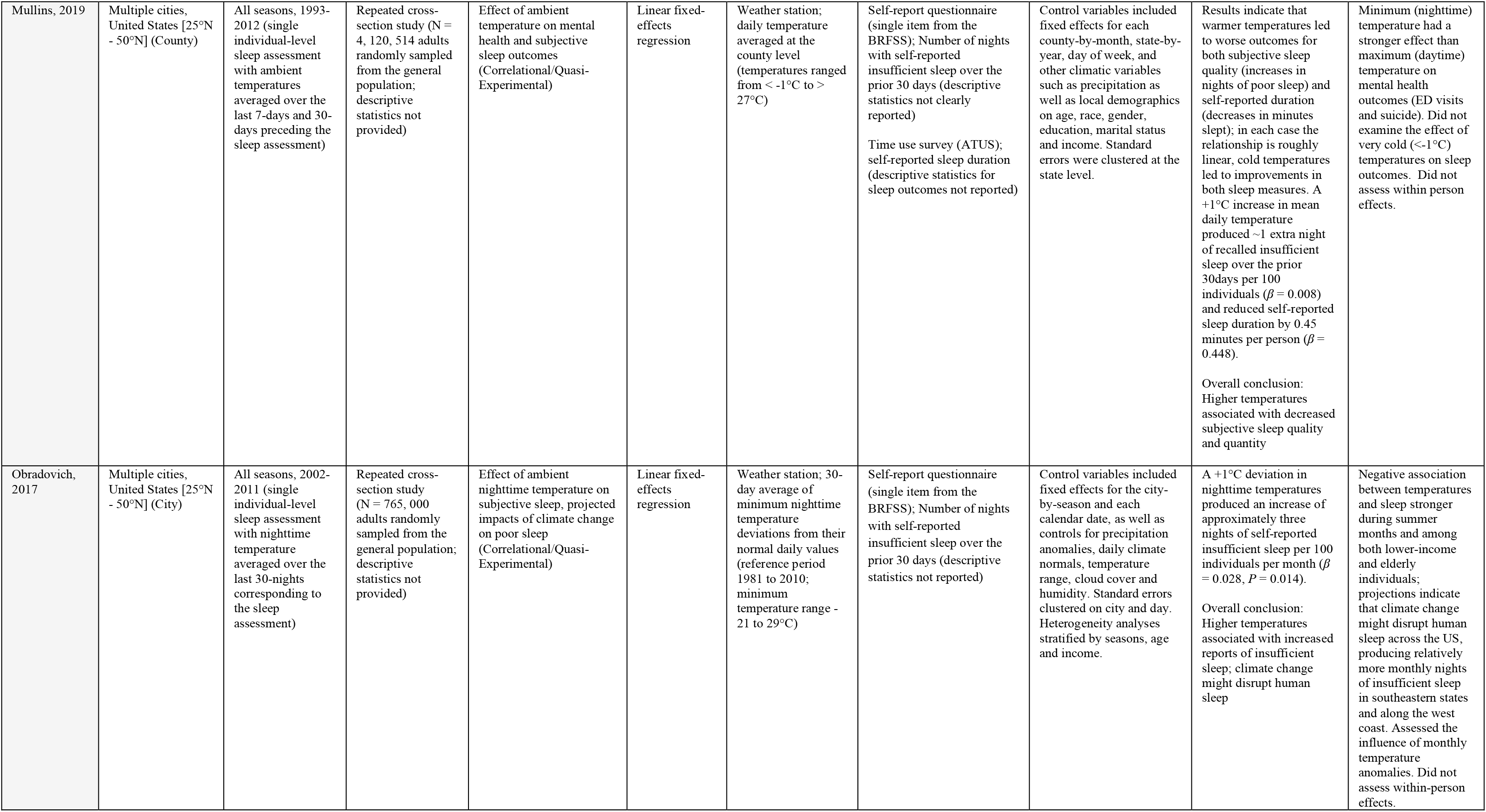
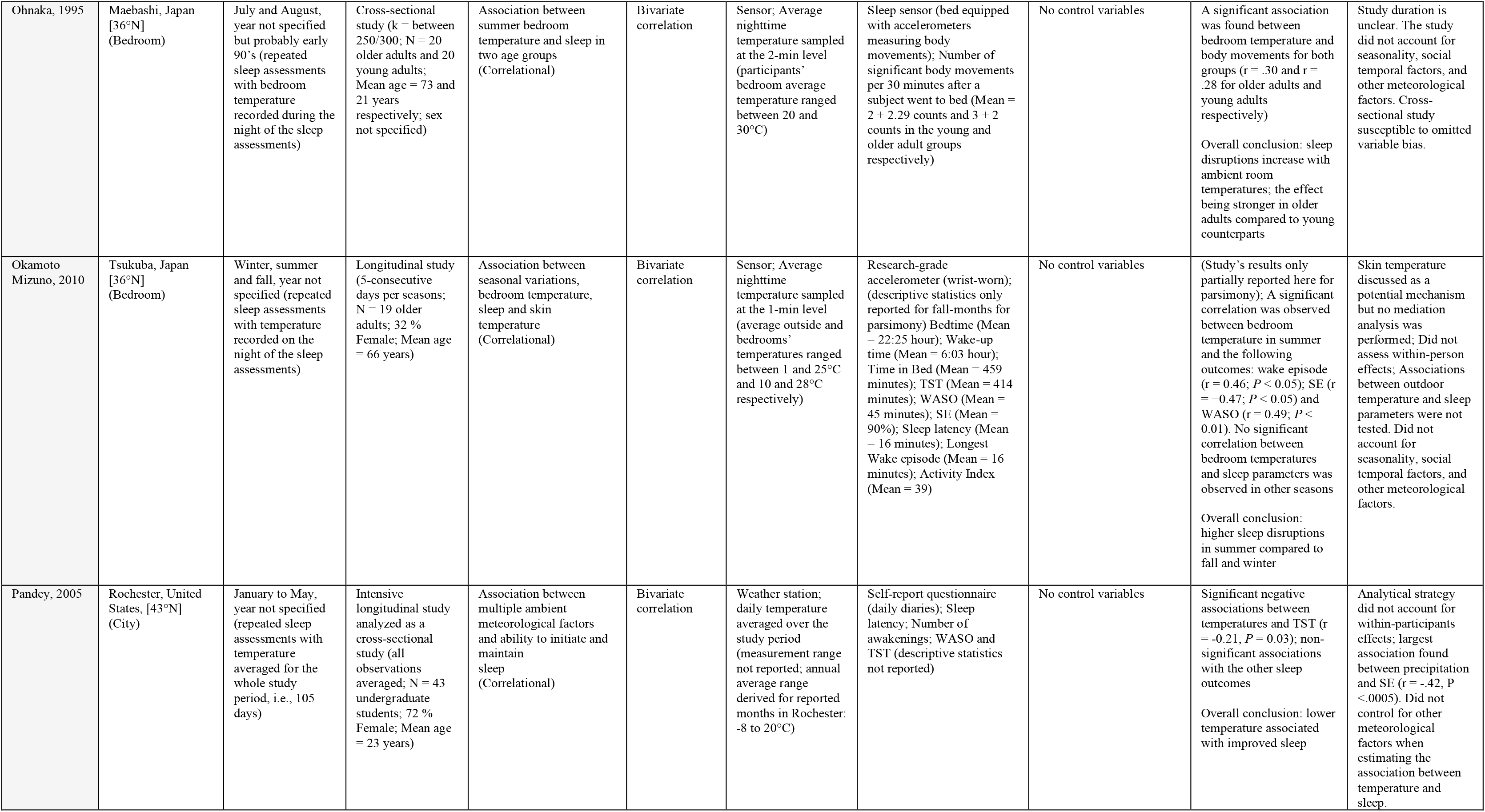
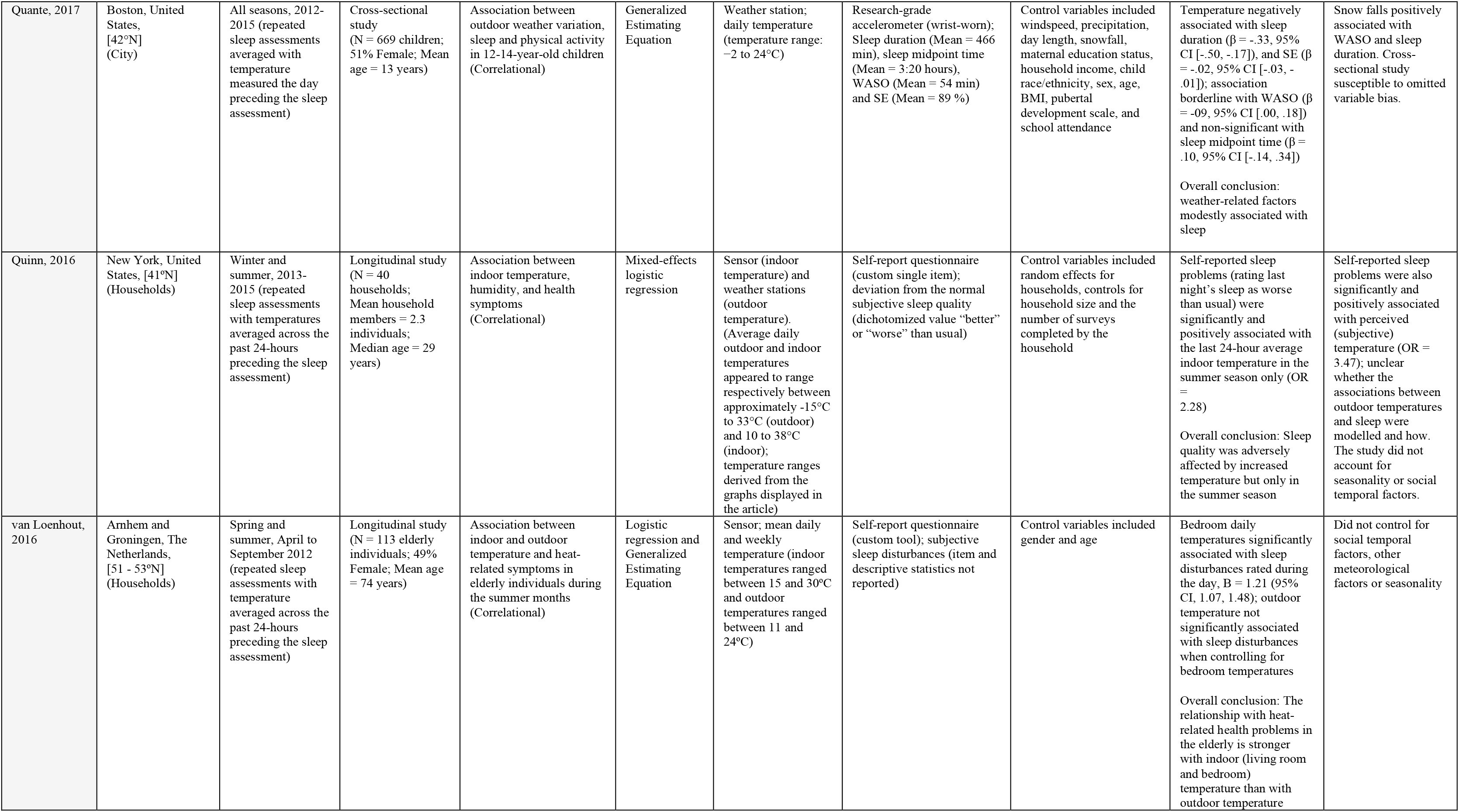
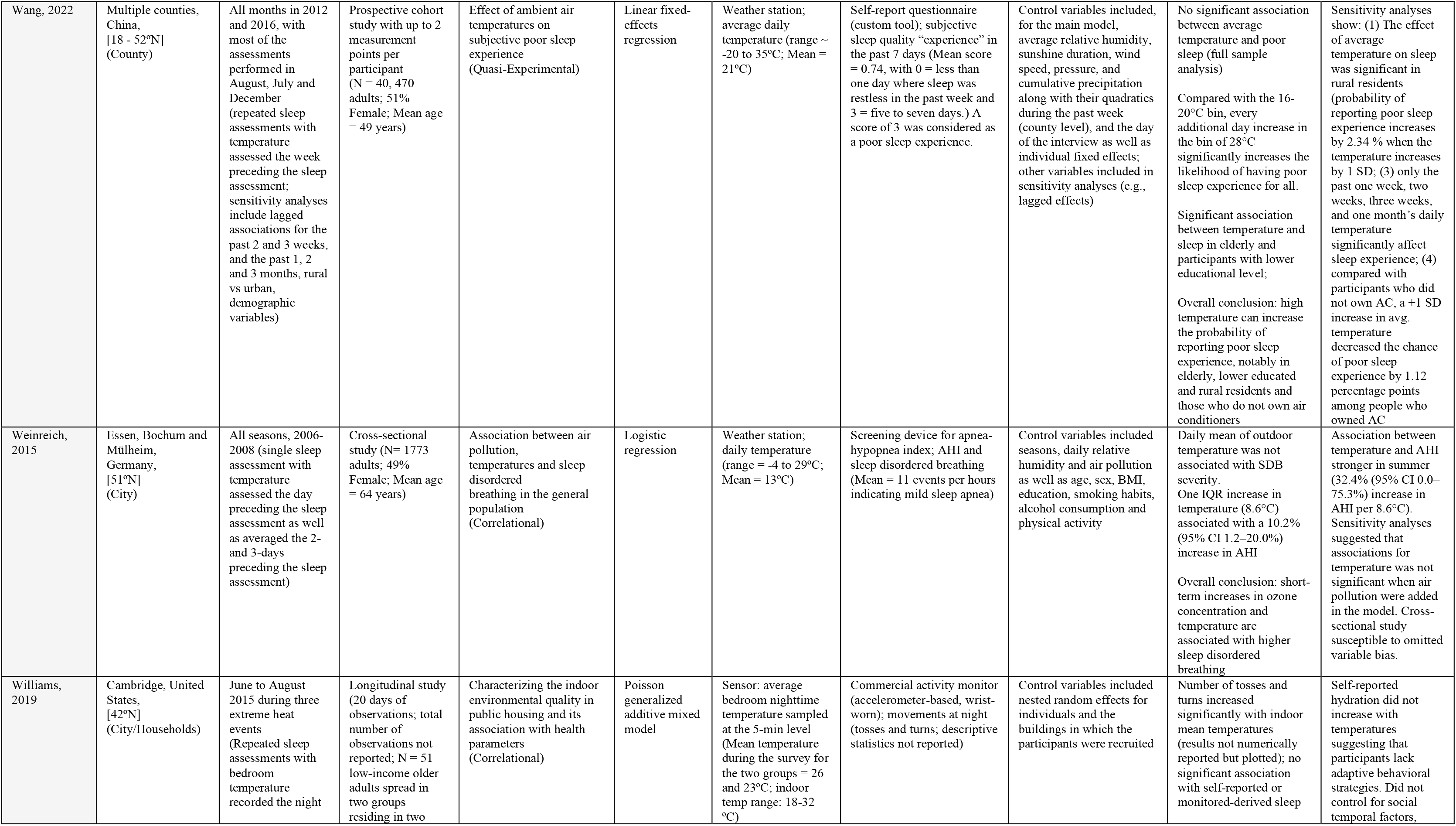
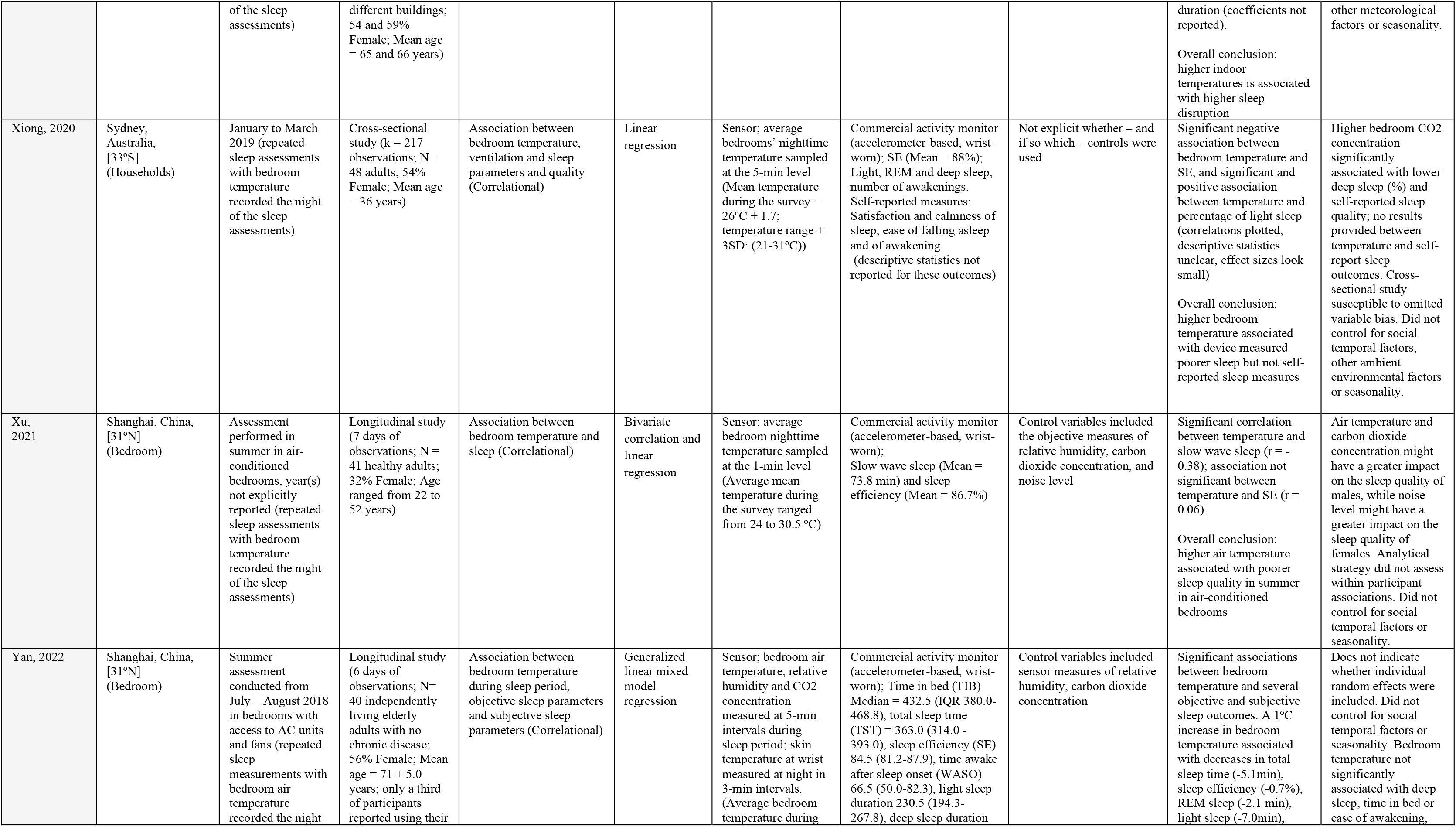
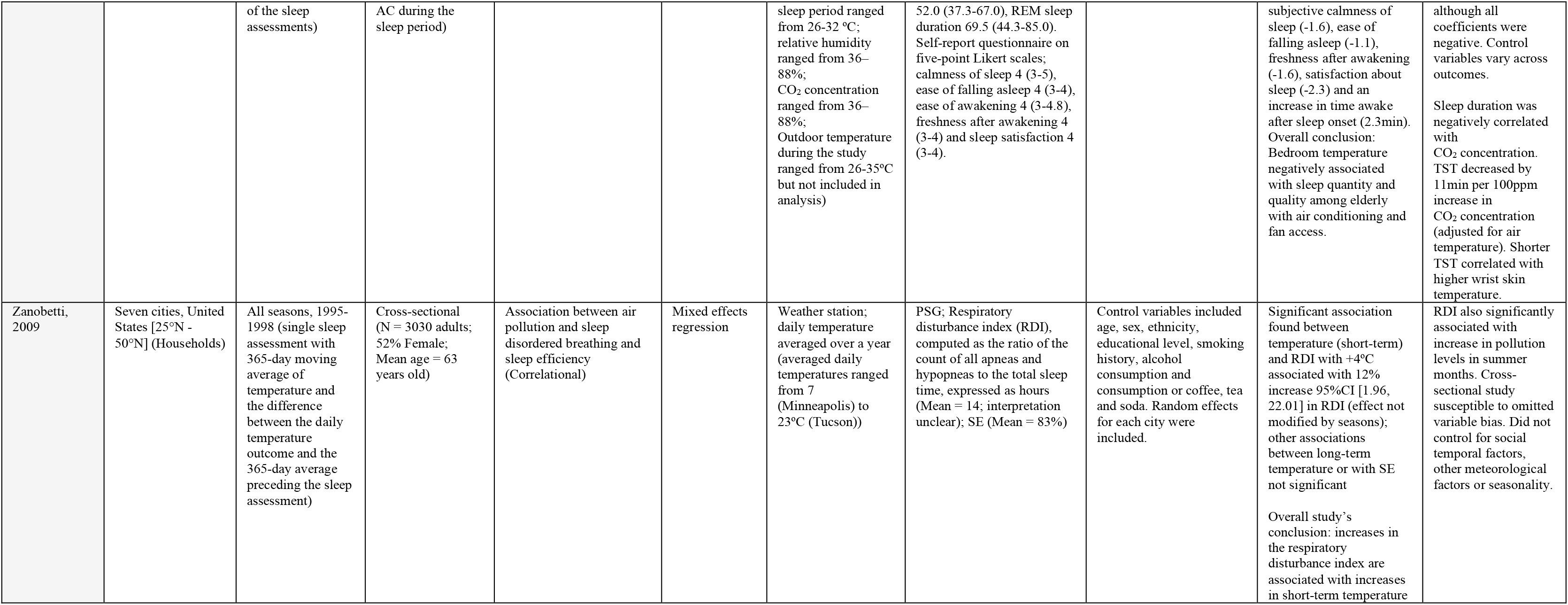
Detailed summary of the studies included in the systematic review

Figure 3 illustrates the range of observed ambient temperatures and latitudes for each of the studies in this review. Aggregating these ranges suggests a greater observational density for hotter compared to colder temperatures (bottom heat bar, Figure 3A) and for northern latitudes compared to both the equatorial region and the southern hemisphere (right heat bar, Figure 3B). Notably, just four (15%) of the studies investigated temperature-related sleep responses in the Tropics, even though the region is home to approximately 40% of the global human population (blue histogram, Figure 3B).^24,61,65,69^

**Figure 3.**
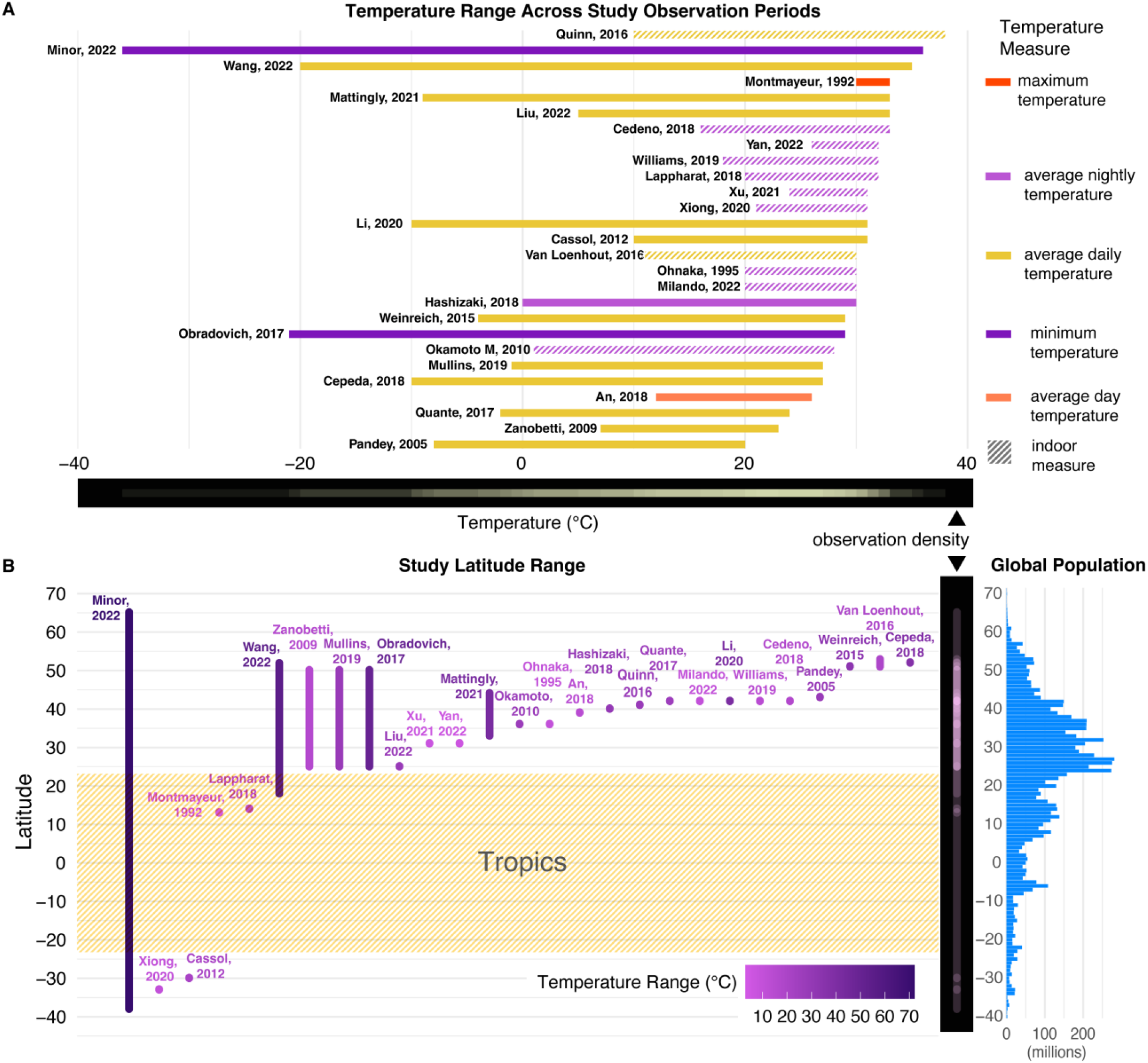
Range of observed ambient temperatures (A) and latitudes (B). of included studies

### Narrative synthesis for the association between ambient temperature and sleep

Overall, most studies (i.e., 22 articles, 80%) concluded that higher temperatures were associated with poorer sleep, with two articles explicitly characterizing the association as “modest” in terms of effect size.^64,79^ Among the five remaining studies: (*i*) two observed a positive effect of temperature on sleep, with one study performed in China reporting a positive association between temperature and self-reported sleep duration,^68^ another, performed in Brazil, showing that apnea-hypopnea index was inversely correlated with ambient temperature;^77^ (*ii*) two other studies were not explicit in their conclusion about this specific temperature-sleep association, with one not observing a significant association between temperature and sleep duration (but delayed sleep timing),^62^ and another observing a significant association between temperature and the number of awakenings but focusing on sleep adaptation in expatriates (not the impact of heat);^61^ and (*iii*) one study showing a non-significant association (neither positive nor negative) between indoor temperature and sleep duration.^75^ In regards to sleep apnea specifically, the abovementioned result^77^ contradicts two other studies included in the present review showing a significant and positive association between temperature and apnea-hypopnea indexes;^76,78^ while a fourth study did not observe significant relationships between temperature (indoor) and apnea-hypopnea indexes.^69^

Results are less contrasted with other sleep outcomes and assessment methods. For example, the four studies using research-grade accelerometers^64,79–81^ all found that higher temperature was associated with poorer sleep (i.e., notably reduced sleep duration and sleep efficiency, as well as increased WASO). In the same vein, four of the six studies using commercial-grade activity monitors found negative associations between ambient temperature and sleep duration,^24,63,71^ efficiency^71,73^ (but see^74,75^ for a non-significant association), WASO and REM sleep,^71^ nighttime body-movements^72^ and slow wave sleep.^74^ Eight studies using questionnaires reported that higher temperature was associated with poorer subjective sleep.^33,34,65–67,69–71^ One study did not observe a significant association between temperature and subjective sleep in adults with episodic migraine,^64^ and another study reported a positive association between temperature and sleep duration in a large cohort of students.^68^ Among the five higher quality studies that fulfilled at least half of the quality criteria,^24,33,62,64,65^ all concluded that higher temperature was associated with poorer sleep outcomes.

For the six studies that identified significant negative effects of temperature on sleep duration, estimated effect sizes appeared to vary in magnitude from modest to large, with effects scaling across the temperature distribution.^24,34,63,67,71,79,80^ For instance, a multi-country study found that at the colder end of local temperature distributions, a +1°C increase reduced sleep by just 0.20 minutes during winter months but sleep was reduced by 0.97 minutes per +1°C during the last month of summer,^24^ whereas a US-based national time use diary study conducted across all seasons estimated a linear sleep reduction of 0.45 minutes per +1°C.^34^ By comparison, a Boston-based study conducted during a summer heatwave estimated a larger sleep reduction of 2.7 minutes per +1°C increase in bedroom temperature for a sample of young adults,^63^ while a Shanghai-based study estimated a large magnitude reduction of 5.10 minutes in total sleep time per +1°C increase for a sample of elderly participants.^71^ Taken together, these results suggest that ambient temperatures may exact both cumulatively and progressively larger sleep impacts at higher ambient temperatures, with one study^24^ estimating that nights exceeding 25°C push 4,600 additional people to experience a short (<7 hour) night of sleep per 100,000 adults compared to the cold optimum identified.

### Other relevant results

Other notable results that further contextualize the temperature-sleep association include (*i*) the role of seasons, (*ii*) the impact of other weather and environmental variables, (*iii*) the presence of non-linear associations between temperature and sleep, (*iv*) the potential mechanisms explaining how temperature impacts sleep, (*v*) the specific role of indoor temperature beyond outdoor temperature, and (*vi*) the role of adaptation measures.

### Seasonality

In regards to seasonality, several studies showed that the negative impact of ambient temperature on sleep was more pronounced in the hottest months of the year as discussed earlier in this review,^24,60,81,82^ although one study failed to replicate this effect.^76^ A study conducted in the Netherlandshighlighted that the seasonality of sleep duration – with people sleeping less during summer and more during winter – appeared to be mainly driven by ambient temperature (i.e., the percentage of variance explained by seasons decreased significantly when temperature was controlled for).^80^ However, another study conducted across the US, found that seasons and day length were the only significant variables associated with sleep duration over 3 principal components mainly representing temperature, wind, humidity and cloud coverage (all of these derived components were not significantly associated with sleep duration).^62^ However, all subjects in this last study were information technology workers working in thermally controlled indoor offices, potentially buffering against outdoor thermal exposures.

### Effect modification with other weather variables

Other weather outcomes such as humidity, precipitation, cloud cover or wind speed were included as control variables in several studies.^24,34,62,68^ In the largest device-based study included in this review,^24^ authors showed that (*i*) the impact of temperature on sleep duration was independent from other weather variables and that (*ii*) higher levels of precipitation, wind speed and cloud coverage marginally increase sleep duration while both low and high levels of humidity significantly reduce sleep duration. Additionally, high diurnal temperature range (the difference between daily maximum and minimum temperature) further reduced sleep duration, albeit to a lesser degree than high night-time temperature.^24^ Concerning humidity specifically, several experimental studies previously showed that the combination of high levels of indoor humidity and heat create the worst conditions for sleep.^32,84^ At high ambient temperature, high levels of humidity compromise the body’s evaporative cooling thermoregulatory response and thus increase the risk of heat stress and hyperthermia, possibly challenging the nocturnal core body temperature decline.^47^ Although evidence remains sparse, one article included in our review investigated the combined effect of heat and humidity on sleep, finding that higher heat index values progressively reduced sleep duration.^24^

### Functional form of the temperature-sleep association

Regarding the functional forms that the temperature-sleep association may take, most studies assumed a linear response but three studies found non-linear associations.^24,33,60^ For example, one global-scale study observed a monotonic decline in sleep duration as night-time temperature increased, but uncovered an inflection point (i.e., a steeper decline) at 10°C, with progressively larger effects at higher temperatures.^24^ Interestingly, the same study found that the marginal effect of ambient temperature on sleep loss was over twice as large in the warmest climate regions compared to the coldest areas, consistent with the kinked functional form identified. A second study found a U-shape association between nighttime temperature and wake after sleep onset, as well as an inverse U-shape functional form for sleep efficiency, with the lowest level of wake after sleep onset and higher sleep efficiency in the range 10-15°C.^60^

### Potential mechanisms

Several researchers have discussed putative causal mechanisms linking temperature and sleep or the role of sleep as a mediator of the association between heat and other health and behavioral outcomes,^14,15,85,86^ although evidence remains limited in this regard. Two studies inferred that body skin temperature may be one of the mechanisms linking temperature and sleep outcomes in the elderly, with higher skin temperature associated with poorer sleep in studies conducted in Japan and China.^71,84^ A second study investigated the interplay between temperature, sleep and mental health.^34^ This last study proposed that sleep loss may be a plausible mechanism explaining the effect of higher temperature on emergency department visits for mental disorders and suicide attempts – both of which exhibited consistent functional forms – but the authors did not perform a proper mediation analysis to test this hypothesis. Another study showed that shorter sleep duration induced by higher temperature may be a mediating factor of the negative association between temperature and cognitive functions.^63^ However, the mediation analysis was only significant for one of five cognitive outcomes assessed. The authors concluded that a small sample size precluded their analysis from yielding conclusive results about the mediating role of sleep in cognitive effects.

### Indoor and outdoor temperatures

Only two studies included in the systematic review combined measures and simultaneous analyses of indoor ambient temperature in parallel with outdoor temperature.^66,70^ A first study, performed in The Netherlands, showed that outdoor temperature was no longer associated with self-reported sleep disturbances when also including indoor (bedroom) temperature in their model specification, with the latter being significantly associated with sleep disturbances.^66^ Interestingly, the second study, conducted in New York, showed that indoor ambient temperature was systematically higher than outdoor temperature, even in summer and with 92 % of the sample reporting air conditioning ownership.^70^ However, this study focused on the effect of indoor temperature and did not statistically control for outdoor temperature when testing the association with sleep, rendering it impossible to disentangle the effect of indoor *versus* outdoor temperature on sleep. A third study measured both indoor and outdoor temperature for elderly residents in Shanghai but only included indoor temperatures in the final analysis.^71^ The range of reported indoor temperatures closely approximated the outdoor range. A separate study found greater sleep loss on days with larger diurnal temperature ranges and cumulatively larger lagged negative effects of outdoor ambient temperature on sleep loss, suggesting that interior environments may trap ambient heat and prolong temperature-related sleep loss.^24^

### Adaptation measures

Finally, only a minority of studies investigated behavioral or technological adaptations that might protect sleep from heat. One study, conducted in Sydney, Australia, showed that air conditioning was rarely operated compared to open windows and the use of fans, and that air conditioning did not interact with the study results showing that higher bedroom temperature was associated with poorer sleep.^73^ Similarly, a separate study found that only a third of elderly participants activated their AC units to cool their rooms at night.^71^ Despite uniform AC and fan access, higher bedroom temperatures were associated with large reductions in sleep quantity and quality. Another study showed that older adults did not report higher levels of hydration (i.e., drinking episodes) when temperature increased, suggesting that participants lacked adaptive behavioral strategies.^72^ One study also investigated whether people adapt to night-time sleep impacts with compensatory sleep during the day (napping), week (catch up sleep) or across summer months (intra-annual acclimatization), but did not find any evidence of sleep adaptation.^24^ This same study also found that residents already living in warmer climate regions were more affected per degree of temperature increase than those living in colder areas, suggestive of limited long-run adaptation. This may indicate an upper threshold for human physiology and appears similar to the pattern observed for the temperature-mortality relationship in Europe.^43^

### Narrative synthesis for available climate change projections

Two studies investigated whether warming nighttime temperatures due to climate change would increase the incidence of insufficient sleep in the future.^24,33^ Obradovich et al. 2017 calculated nighttime temperature anomalies for 2050 and 2099 for the Representative Concentration Pathways “high greenhouse gas (GHG) concentration” scenario (RCP8.5; IPCC) and the United States based on a large empirical self-reported sleep dataset.^33^ Assuming no further adaptation and that the same functional sleep response persists in the future climate, these authors inferred that climate change may cause between 6 to 14 additional nights of insufficient sleep per 100 individuals by 2050 and 2099, with the greatest increase in climate change-induced nights of insufficient sleep evident in areas of the western and northern United States. Assuming that future adaptation responses do not exceed those observed across the diverse global climate regions examined in the recent historical record, Minor et al. 2022 projected the impact of climate change on sleep for two scenarios: the end-of the century GHG stabilization scenario (RCP4.5) and the increasing GHG concentration scenario (RCP8.5), using empirical sleep data from 68 countries.^24^ Their globally averaged, population-weighted projections indicate that by 2099, sleep loss might vary between approximately 50 hours per year in a stabilized GHG scenario (RCP4.5) to 58 hours under a less plausible increasing GHG scenario (RCP8.5). By the end of the century, the authors separately estimate that individuals might experience 13 (RCP4.5) to 15 (RCP8.5) excess short (<7 hours) nights of sleep per person per year. These last simulations also indicate that global inequalities in the effect of climate change on sleep loss may scale with future greenhouse gas concentrations, with the warmest regions of the world disproportionately impacted. The authors reported that future sleep loss may also be larger for certain demographics that were less represented in their sample composition, referring to their subgroup analyses that found larger marginal effects for residents from lower-income countries (by a factor of approximately 3), older adults (by a factor of 2) and women (∼25% higher).^24^

## Discussion

Projection studies estimate that, with ongoing climate change, the number of nights with insufficient sleep may significantly increase by the end of the century.^24,33^ Since the global prevalence of poor sleep is already high, it is crucial to develop a detailed and comprehensive understanding of the effect of temperature on sleep. The present systematic review, which includes 27 original articles, shows that higher outdoor or indoor ambient temperatures, expressed either as daily mean or nighttime temperature, are negatively associated with various sleep outcomes worldwide. This negative effect of higher ambient temperatures on sleep is stronger in the warmest months of the year, among vulnerable populations, notably in the elderly, and in the warmest areas of the world. This result seems consistent across various sleep indicators including sleep quantity, timing or quality and measured via various means including questionnaires, polysomnography, research-grade or commercial activity monitors (see Tables 2 and 3 for a summary of the results). Although the heat-related results are in accordance with those from previous reviews focused on experimental studies manipulating indoor temperatures,^26^ studies investigating both cold and hot outdoor ambient temperatures have found elevated sleep duration during colder temperatures, suggesting that people may be better at adapting to low ambient temperature than to ambient heat.

Nonetheless, the methodological quality of most studies included in the present systematic review is low (see Table 4). The current literature is notably limited by a relatively poor consideration of key potential individual, spatiotemporal and social confounders. For instance, only 41% (11/27) of the studies statistically adjusted for location-specific seasonality (see Table 4), even though seasonality is associated with changes in daylight, environmental characteristics and behaviors that may also influence or otherwise spuriously associate with sleep. Although the impact of temperature on sleep appears robust – even when controlling for other weather variables (i.e., precipitation, cloud cover, humidity, wind speed, diurnal temperature range), only few studies properly handle these covariates.^24,34,62^ The negative association between ambient temperature and sleep also remains significant when controlling for adaptation measures such as the utilization of air conditioning, but this should also be further explored.^34,65,70,71^ Additionally, only 18% (5/27) of the studies assessed the plausible lagged effects of ambient temperature conditions on sleep in addition to the contemporaneous effect (see Table 4).^24,61,64,82^

**Table 4.** Methodological quality of included studies As shown in Table 4, study quality was generally low as assessed by the 14-item research quality checklist developed for this systematic review (i.e., the quality criteria were only met by ∼30% of all cells, marked in the table in green). Although there was a high degree of heterogeneity between studies in terms of evaluated quality, some specific criteria were rarely met by the extant literature such as the combined measurement and analysis of both indoor and outdoor temperatures (estimated by only one prior study), the inclusion of personal cooling strategies (e.g., fans), or other time-varying meteorological controls that might otherwise confound inference between temperature and sleep.

|  | Item 1:<br>non-linearity<br>inspected | Item 2:<br>exposure<br>assessed<br>more<br>than once | Item 3:<br>within-participant<br>variation<br>analyzed | Item 4:<br>sleep and<br>temperatures<br>measured over<br>the whole year | Item 5:<br>seasonality<br>or day<br>length<br>controlled | Item 6:<br>temporal<br>variables<br>controlled | Item 7:<br>humidity<br>controlled | Item 8:<br>wind speed<br>controlled | Item 9:<br>cloud cover<br>controlled | Item 10:<br>precipitation<br>controlled | Item 11:<br>indoor and<br>outdoor<br>temperature<br>measured and<br>analyzed | Item 12:<br>lagged<br>temperature<br>effects<br>inspected | Item 13:<br>personal<br>cooling<br>technologies<br>measured and<br>analyzed | Item 14: sleep<br>measured with<br>questionnaires<br>and devices |
| --- | --- | --- | --- | --- | --- | --- | --- | --- | --- | --- | --- | --- | --- | --- |
| An, 2018 | 0 | 1 | 1 | 0 | 0 | 0 | 0 | 1 | 0 | 1 | 0 | 0 | 0 | 0 |
| Cassol, 2012 | 0 | 0 | 0 | 1 | 1 | 0 | 0 | 0 | 0 | 0 | 0 | 0 | 0 | 0 |
| Cedeño L, 2018 | 0 | 1 | 1 | 0 | 0 | 0 | 0 | 0 | 0 | 0 | 0 | 0 | 1 | 0 |
| Cepeda, 2018 | 0 | 1 | 1 | 1 | 1 | 1 | 0 | 0 | 0 | 0 | 0 | 0 | 0 | 0 |
| Hashizaki, 2018 | 1 | 1 | 1 | 1 | 1 | 1 | 0 | 0 | 0 | 0 | 0 | 0 | 0 | 0 |
| Lappharat, 2018 | 0 | 0 | 0 | 0 | 0 | 0 | 0 | 0 | 0 | 0 | 0 | 0 | 0 | 1 |
| Li, 2020 | 1 | 1 | 1 | 1 | 0 | 1 | 0 | 0 | 0 | 0 | 0 | 1 | 0 | 1 |
| Liu, 2022 | 0 | 0 | 0 | 1 | 0 | 0 | 0 | 0 | 0 | 0 | 0 | 0 | 0 | 0 |
| Mattingly, 2021 | 0 | 1 | 1 | 1 | 1 | 0 | 1 | 1 | 1 | 0 | 0 | 0 | 0 | 0 |
| Milando, 2022 | 0 | 1 | 0 | 0 | 0 | 1 | 0 | 0 | 0 | 0 | 0 | 0 | 0 | 1 |
| Minor, 2022 | 1 | 1 | 1 | 1 | 1 | 1 | 1 | 1 | 1 | 1 | 0 | 1 | 0 | 0 |
| Montmayer, 1992 | 0 | 1 | 0 | 0 | 0 | 0 | 0 | 0 | 0 | 0 | 0 | 1 | 0 | 0 |
| Mullins, 2019 | 1 | 0 | 0 | 1 | 1 | 1 | 1 | 0 | 0 | 1 | 0 | 0 | 0 | 0 |
| Obradovich, 2017 | 1 | 0 | 0 | 1 | 1 | 1 | 1 | 0 | 1 | 1 | 0 | 0 | 0 | 0 |
| Ohnaka, 1995 | 0 | 1 | 0 | 0 | 0 | 0 | 0 | 0 | 0 | 0 | 0 | 0 | 0 | 0 |
| Okamoto M, 2010 | 0 | 1 | 0 | 0 | 0 | 0 | 0 | 0 | 0 | 0 | 0 | 0 | 0 | 0 |
| Pandey, 2005 | 0 | 0 | 0 | 0 | 0 | 0 | 0 | 0 | 0 | 0 | 0 | 0 | 0 | 0 |
| Quante, 2017 | 1 | 0 | 0 | 1 | 1 | 0 | 0 | 1 | 0 | 1 | 0 | 0 | 0 | 1 |
| Quinn, 2016 | 0 | 1 | 1 | 1 | 0 | 0 | 0 | 0 | 0 | 0 | 0 | 0 | 0 | 0 |
| Van Loenhout, 2016 | 0 | 1 | 1 | 0 | 0 | 0 | 0 | 0 | 0 | 0 | 1 | 0 | 0 | 0 |
| Wang, 2022 | 1 | 1 | 1 | 0 | 1 | 1 | 1 | 1 | 0 | 1 | 0 | 1 | 1 | 0 |
| Weinreich, 2015 | 1 | 0 | 0 | 0 | 1 | 0 | 1 | 0 | 0 | 0 | 0 | 1 | 0 | 0 |
| Williams, 2019 | 1 | 1 | 1 | 0 | 0 | 0 | 0 | 0 | 0 | 0 | 0 | 0 | 1 | 1 |
| Xiong, 2020 | 0 | 1 | 0 | 0 | 0 | 0 | 0 | 0 | 0 | 0 | 0 | 0 | 0 | 1 |
| Xu, 2021 | 0 | 1 | 0 | 0 | 0 | 0 | 1 | 0 | 0 | 0 | 0 | 0 | 0 | 1 |
| Yan, 2022 | 0 | 1 | 0 | 0 | 0 | 0 | 1 | 0 | 0 | 0 | 0 | 0 | 1 | 1 |
| Zanobetti, 2009 | 0 | 0 | 0 | 1 | 1 | 0 | 1 | 1 | 1 | 1 | 0 | 0 | 0 | 0 |

Moreover, the relative importance of indoor and outdoor ambient temperatures remains remarkably unclear and virtually unassessed. According to the only study that accounted for both measures, indoor temperature (i.e., measured in the bedroom), more than outdoor temperature, appeared to drive the relationship between ambient heat and sleep.^66^ It is worth highlighting that climate change is shifting the underlying distribution of local outdoor temperatures, yet adaptation will continue to transpire both outdoors and indoors. Thus, both ambient temperature measures likely impact human sleep through potentially distinct and/or overlapping pathways that should be investigated in future research, both independently – and where possible – in combination. Similarly, it’s unclear how daytime and nighttime temperatures interact to impact sleep. To our knowledge, only one study tested this effect and found that a higher diurnal temperature range was independently associated with decreased sleep duration.^24^

These limitations and the quality assessment performed for this review help to draw a set of recommendations for future studies. First, researchers and funding agencies should pursue large-scale cooperative projects leveraging repeated person-level sleep measures (including, but not limited to personal sensing technologies) and longitudinal study designs across larger, and more globally diverse populations, and for longer periods of unobtrusive observation.^87^ The geographic distribution of studies conducted so far does not cover the global distribution of the human population (Figures 2, 3B). Drawing on these timeseries data, researchers should explicitly control for time-invariant between-individual differences to identify within-person temperature-sleep responses.

Second, key spatiotemporally-varying factors should be more consistently controlled in future studies, including daily precipitation, percentage of cloud cover, relative humidity, average wind speed, local climatological conditions for these meteorological variables, and potentially, other relevant ambient environmental factors (e.g., air pollution, day length, etc.).^24^ Further, for quasi-experimental study designs that seek to identify plausibly causal effects from as good as random variation in ambient temperature fluctuations in real-life environments, researchers should control for location-specific seasonality as well as socio-temporal trends by accounting for day of study-specific shocks due to calendar-induced behavioral changes and macro events that might spuriously associate with both temperature and sleep outcomes.

Third, future studies should strive to investigate the effects of indoor *versus* outdoor temperatures and diurnal *versus* nighttime temperatures on sleep.^66^ Fourth, studies should consider the lagged and cumulative effects of temperature and other meteorological variables on sleep outcomes. Fifth, behavioral and technological adaptation measures should be more consistently measured and included in analyses; this includes hydration, behaviors related to sleep hygiene and the utilization of fans or air conditioners.^31,63,72^ Sixth, non-linearity in the association between temperature and sleep, as well as other meteorological controls, should be systematically inspected and reported.^24^ Seventh, a priority should be given to vulnerable populations who received scant attention so far, including habitants of low-income countries, individuals with low financial resources within high-income countries, women in peripartum period,^88^ developing infants and children,^79,89^ residents living in the tropics (Figure 3B), residents living in extremely cold and hot environments (Figure 3A), incarcerated populations with limited environmental controls, individuals with mental health disorders, and those with sleep disorders such as insomnia and restless legs syndrome.^54^ Eighth, and as argued before,^15,85^ more mechanistic studies are still very much needed to both better understand (*i*) the potential mediators of the temperature-sleep association beyond physiological parameters (e.g., mental health)^9^, and (*ii*), although not the main focus of this systematic review, the contribution of sleep issues in the pathway between ambient temperatures and health outcomes (e.g., mortality).^29,90^ Given the congruence between the recently identified temperature-sleep functional response and temperature-mental health functional forms,^11,24,33,34,91^ carefully designed field experiments that enable rigorous assessments of mediation while also experimentally shutting down other temperature-sensitive pathways are needed to inform well-targeted policy responses that bolster heat and sleep resilience. Finally, only 30% (8/27) of the studies in this review investigated temperature-sleep relationships across multiple cities or geographic regions,^24,33,34,62,65,66,76,82^ and only one featured multiple countries.^24^ Since spatial autocorrelation is likely high within single county and city studies – likely introducing bias to results and interpretation^92^ – researchers should strive to carry out research investigations and multi-country collaborations across diverse geographic regions while statistically accounting for spatially correlated errors.

### Fostering adaptation

Beyond observational studies, there is an urgent need for interventional studies aiming to foster heat adaptation at different levels, from interventions focused on individuals to environmental and structural modifications.^93^ At the individual level, evidence-based sleep hygiene measures should be tested to see whether such behavioral measures can foster adaptation to ambient heat. This includes general sleep health measures, such as the avoidance of caffeine, nicotine, alcohol and daytime naps, stress management, sleep timing regularity, management of bedroom noise and artificial light.^94^ Additionally, heat-specific behavioral adaptations should also be assessed, including cool showers before bedtime, the use of fans (when relative humidity <30%),^95,96^ water sprays, daytime hydration, reduced bedding, and light cotton clothing.^88^ Traditional lifestyle interventions, such as the promotion of regular physical activity, are also crucial given the role of physical fitness towards heat adaptation.^97,98^ These interventions could be implemented through traditional randomized controlled trials or using innovative designs such as just-in-time interventions using weather forecasts that might be particularly relevant for heatwaves.^99,100^ At the societal level, equitable adaptation should be promoted^101^. These efforts should ideally be combined with environmental measures such as urban greening,^102^ urban water features, passive cooling and the improvement of buildings’ insulation and ventilation systems.^31,103^

## Conclusion

The present systematic review shows that higher temperatures are generally associated with poorer sleep outcomes worldwide. Given the absence of solid evidence on fast sleep adaptation to heat, rising average temperatures induced by climate change pose a serious threat to human sleep and therefore human health, performance, and wellbeing. Although this work identified several methodological limitations of the extant literature, a strong body of evidence from both this systematic review and previous experimental studies converge on the negative impact of elevated temperatures on sleep quality and quantity. Pertinent to policymakers, planners and sleep researchers, the intensity of night-time warming is projected to continue to exceed daytime warming in most populated areas,^29,104–106^ while urbanization will likely further exacerbate night-time ambient heat exposure for most of humanity.^107^ Even if these relationships and their associated pathways can be refined further through future well-designed observational studies as we advise here, we argue that interventional studies are now urgently needed to foster adaptation and safeguard the essential restorative role of sleep in a hotter world.

## Data Availability

Not appropriate

## Acknowledgements

The authors want to thanks Pr Manolis Kogevinas and Dr Michael P Mead for their feedbacks on the manuscript.

## References

1. Whitmee S, Haines A, Beyrer C, Boltz F, Capon AG, Dias BF de S, et al. Safeguarding human health in the Anthropocene epoch: report of The Rockefeller Foundation–Lancet Commission on planetary health. The Lancet. 2015 Nov;386(10007):1973–2028.

2. Ebi KL, Frumkin H, Hess JJ. Protecting and promoting population health in the context of climate and other global environmental changes. Anthropocene. 2017 Sep 1;19:1–12.

3. Ebi KL, Capon A, Berry P, Broderick C, Dear R de, Havenith G, et al. Hot weather and heat extremes: health risks. The Lancet. 2021 Aug 21;398(10301):698–708.

4. Romanello M, McGushin A, Napoli CD, Drummond P, Hughes N, Jamart L, et al. The 2021 report of the Lancet Countdown on health and climate change: code red for a healthy future. The Lancet. 2021 Oct 30;398(10311):1619–62.

5. Watts N, Amann M, Arnell N, Ayeb-Karlsson S, Belesova K, Boykoff M, et al. The 2019 report of The Lancet Countdown on health and climate change: ensuring that the health of a child born today is not defined by a changing climate. The Lancet. 2019 Nov 16;394(10211):1836–78.

6. Lee H, Myung W, Kim H, Lee EM, Kim H. Association between ambient temperature and injury by intentions and mechanisms: A case-crossover design with a distributed lag nonlinear model. Science of The Total Environment. 2020 Dec 1;746:141261.

7. Parks RM, Bennett JE, Tamura-Wicks H, Kontis V, Toumi R, Danaei G, et al. Anomalously warm temperatures are associated with increased injury deaths. Nat Med. 2020 Jan;26(1):65–70.

8. Onozuka D, Hagihara A. All-Cause and Cause-Specific Risk of Emergency Transport Attributable to Temperature: A Nationwide Study. Medicine (Baltimore). 2015 Dec;94(51):e2259.

9. Thompson R, Hornigold R, Page L, Waite T. Associations between high ambient temperatures and heat waves with mental health outcomes: a systematic review. Public Health. 2018 Aug 1;161:171–91.

10. Liu Y, Saha S, Hoppe BO, Convertino M. Degrees and dollars – Health costs associated with suboptimal ambient temperature exposure. Science of The Total Environment. 2019 Aug 15;678:702–11.

11. Park RJ, Behrer AP, Goodman J. Learning is inhibited by heat exposure, both internationally and within the United States. Nat Hum Behav. 2021 Jan;5(1):19–27.

12. Dasgupta S, Maanen N van, Gosling SN, Piontek F, Otto C, Schleussner CF. Effects of climate change on combined labour productivity and supply: an empirical, multi-model study. The Lancet Planetary Health. 2021 Jul 1;5(7):e455–65.

13. Adélaïde L, Chanel O, Pascal M. Health effects from heat waves in France: an economic evaluation. Eur J Health Econ. 2022 Feb 1;23(1):119–31.

14. Rifkin DI, Long MW, Perry MJ. Climate change and sleep: A systematic review of the literature and conceptual framework. Sleep Med Rev. 2018 Dec;42:3–9.

15. Obradovich N, Migliorini R. Sleep and the human impacts of climate change. Sleep Medicine Reviews. 2018 Dec 1;42:1–2.

16. Chevance G, Fresán U, Hekler E, Edmondson D, Lloyd SJ, Ballester J, et al. Thinking Health-related Behaviors in a Climate Change Context: A Narrative Review. Ann Behav Med. 2022 Jul 21;kaac039.

17. Knutson KL, Van Cauter E, Rathouz PJ, Yan LL, Hulley SB, Liu K, et al. Association Between Sleep and Blood Pressure in Midlife: The CARDIA Sleep Study. Archives of Internal Medicine. 2009 Jun 8;169(11):1055–61.

18. Knutson KL, Van Cauter E. Associations between sleep loss and increased risk of obesity and diabetes. Ann N Y Acad Sci. 2008;1129:287–304.

19. Kakizaki M, Inoue K, Kuriyama S, Sone T, Matsuda-Ohmori K, Nakaya N, et al. Sleep duration and the risk of prostate cancer: the Ohsaki Cohort Study. Br J Cancer. 2008 Jul 8;99(1):176–8.

20. Bernert RA, Kim JS, Iwata NG, Perlis ML. Sleep Disturbances as an Evidence-Based Suicide Risk Factor. Curr Psychiatry Rep. 2015 Feb 21;17(3):15.

21. Williamson A, Lombardi DA, Folkard S, Stutts J, Courtney TK, Connor JL. The link between fatigue and safety. Accident Analysis & Prevention. 2011 Mar 1;43(2):498– 515.

22. Chepesiuk R. Missing the Dark: Health Effects of Light Pollution. Environ Health Perspect. 2009 Jan;117(1):A20–7.

23. Smith MG, Cordoza M, Basner M. Environmental Noise and Effects on Sleep: An Update to the WHO Systematic Review and Meta-Analysis. Environmental Health Perspectives. 130(7):076001.

24. Minor K, Bjerre-Nielsen A, Jonasdottir SS, Lehmann S, Obradovich N. Rising temperatures erode human sleep globally. One Earth. 2022 May 20;5(5):534–49.

25. Harding EC, Franks NP, Wisden W. The Temperature Dependence of Sleep. Frontiers in Neuroscience. 2019 Feb 24;13: 336.

26. Buguet A. Sleep under extreme environments: effects of heat and cold exposure, altitude, hyperbaric pressure and microgravity in space. Journal of the neurological sciences. 2007 Nov;262(1–2):145–52.

27. Krauchi K, Deboer T. The interrelationship between sleep regulation and thermoregulation. Frontiers in bioscience. 2010 Jan;15:604–25.

28. Seltenrich N. Between Extremes: Health Effects of Heat and Cold. Environmental Health Perspectives. 2015 Nov;123(11):A275–9.

29. He C, Kim H, Hashizume M, Lee W, Honda Y, Kim SE, et al. The effects of night-time warming on mortality burden under future climate change scenarios: a modelling study. The Lancet Planetary Health. 2022 Aug 1;6(8):e648–57.

30. Wu Y, Li S, Zhao Q, Wen B, Gasparrini A, Tong S, et al. Global, regional, and national burden of mortality associated with short-term temperature variability from 2000–19: a three-stage modelling study. The Lancet Planetary Health. 2022 May 1;6(5):e410–21.

31. Jay O, Capon A, Berry P, Broderick C, Dear R de, Havenith G, et al. Reducing the health effects of hot weather and heat extremes: from personal cooling strategies to green cities. The Lancet. 2021 Aug 21;398(10301):709–24.

32. Okamoto-Mizuno K, Mizuno K, Michie S, Maeda A, Iizuka S. Effects of humid heat exposure on human sleep stages and body temperature. Sleep. 1999 Sep 15;22(6):767– 73.

33. Obradovich N, Migliorini R, Mednick SC, Fowler JH. Nighttime temperature and human sleep loss in a changing climate. Sci Adv. 2017 May;3(5):e1601555.

34. Mullins JT, White C. Temperature and mental health: Evidence from the spectrum of mental health outcomes. Journal of Health Economics. 2019;68:102240.

35. Léger D, Poursain B, Neubauer D, Uchiyama M. An international survey of sleeping problems in the general population. Curr Med Res Opin. 2008 Jan;24(1):307–17.

36. Simonelli G, Marshall NS, Grillakis A, Miller CB, Hoyos CM, Glozier N. Sleep health epidemiology in low and middle-income countries: a systematic review and meta-analysis of the prevalence of poor sleep quality and sleep duration. Sleep Health. 2018 Jun;4(3):239–50.

37. Andrijevic M, Byers E, Mastrucci A, Smits J, Fuss S. Future cooling gap in shared socioeconomic pathways. Environ Res Lett. 2021 Sep;16(9):094053.

38. IPCC, 2021: Climate Change 2021: The Physical Science Basis. Contribution of Working Group I to the Sixth Assessment Report of the Intergovernmental Panel on Climate Change [ (eds.)]. Cambridge University Press.

39. Achebak H, Devolder D, Ballester J. Heat-related mortality trends under recent climate warming in Spain: A 36-year observational study. PLOS Medicine. 2018 juil;15(7):e1002617.

40. Vogel MM, Zscheischler J, Wartenburger R, Dee D, Seneviratne SI. Concurrent 2018 Hot Extremes Across Northern Hemisphere Due to Human-Induced Climate Change. Earth’s Future. 2019;7(7):692–703.

41. Thiery W, Lange S, Rogelj J, Schleussner CF, Gudmundsson L, Seneviratne SI, et al. Intergenerational inequities in exposure to climate extremes. Science. 2021 Oct 8;374(6564):158–60.

42. Lee W, Kim Y, Sera F, Gasparrini A, Park R, Choi HM, et al. Projections of excess mortality related to diurnal temperature range under climate change scenarios: a multi-country modelling study. The Lancet Planetary Health. 2020 Nov 1;4(11):e512–21.

43. Martínez-Solanas È, Quijal-Zamorano M, Achebak H, Petrova D, Robine JM, Herrmann FR, et al. Projections of temperature-attributable mortality in Europe: a time series analysis of 147 contiguous regions in 16 countries. The Lancet Planetary Health. 2021 Jul 1;5(7):e446–54.

44. Quijal-Zamorano M, Martínez-Solanas È, Achebak H, Petrova D, Robine JM, Herrmann FR, et al. Seasonality reversal of temperature attributable mortality projections due to previously unobserved extreme heat in Europe. The Lancet Planetary Health. 2021 Sep 1;5(9):e573–5.

45. Lan L, Tsuzuki K, Liu YF, Lian ZW. Thermal environment and sleep quality: A review. Energy and Buildings. 2017 Aug 15;149:101–13.

46. Troynikov O, Watson CG, Nawaz N. Sleep environments and sleep physiology: A review. Journal of Thermal Biology. 2018 Dec 1;78:192–203.

47. Manzar MdD, Sethi M, Hussain ME. Humidity and sleep: a review on thermal aspect. Biological Rhythm Research. 2012;43(4):439–57.

48. Freedman RR, Roehrs TA. Effects of REM sleep and ambient temperature on hot flash-induced sleep disturbance. Menopause (New York, NY). 2006 Aug;13(4):576–83.

49. Karacan I, Thornby JI, Anch AM, Williams RL, Perkins HM. Effects of high ambient temperature on sleep in young men. Aviat Space Environ Med. 1978 Jul;49(7):855–60.

50. Okamoto-Mizuno K, Tsuzuki K, Mizuno K. Effects of mild heat exposure on sleep stages and body temperature in older men. Int J Biometeorol. 2004 Sep;49(1):32–6.

51. Moher D, Shamseer L, Clarke M, Ghersi D, Liberati A, Petticrew M, et al. Preferred reporting items for systematic review and meta-analysis protocols (PRISMA-P) 2015 statement. Systematic Reviews. 2015 Jan 1;4(1):1.

52. Min KB, Lee S, Min JY. High and low ambient temperature at night and the prescription of hypnotics. Sleep. 2021 May;44(5).

53. Hjorth MF, Chaput JP, Michaelsen K, Astrup A, Tetens I, Sjödin A. Seasonal variation in objectively measured physical activity, sedentary time, cardio-respiratory fitness and sleep duration among 8-11 year-old Danish children: a repeated-measures study. BMC Public Health. 2013 Sep 8;13:808.

54. Bjorvatn B, Waage S, Pallesen S. The association between insomnia and bedroom habits and bedroom characteristics: an exploratory cross-sectional study of a representative sample of adults. Sleep Health: Journal of the National Sleep Foundation. 2018 Apr;4(2):188–93.

55. Tsang TW, Mui KW, Wong LT. Investigation of thermal comfort in sleeping environment and its association with sleep quality. Building & Environment. 2021;187:N.PAG-N.PAG.

56. Yetish G, Kaplan H, Gurven M, Wood B, Pontzer H, Manger PR, et al. Natural sleep and its seasonal variations in three pre-industrial societies. Current biology : CB. 2015 Nov;25(21):2862–8.

57. Imagawa H, Rijal HB. Field survey of the thermal comfort, quality of sleep and typical occupant behaviour in the bedrooms of Japanese houses during the hot and humid season. Architectural Science Review. 2015 Jan 2;58(1):11–23.

58. Zhang X, Luo G, Xie J, Liu J. Associations of bedroom air temperature and CO2 concentration with subjective perceptions and sleep quality during transition seasons. Indoor Air. 2021;31(4):1004–17.

59. Study Quality Assessment Tools | NHLBI, NIH [Internet]. [cited 2022 Sep 28]. Available from: https://www.nhlbi.nih.gov/health-topics/study-quality-assessment-tools

60. Hashizaki M, Nakajima H, Shiga T, Tsutsumi M, Kume K. A longitudinal large-scale objective sleep data analysis revealed a seasonal sleep variation in the Japanese population. Chronobiol Int. 2018 Jul;35(7):933–45.

61. Montmayeur A, Buguet A. Sleep patterns of European expatriates in a dry tropical climate. Journal of Sleep Research. 1992;1(3):191–6.

62. Mattingly SM, Grover T, Martinez GJ, Aledavood T, Robles-Granda P, Nies K, et al. The effects of seasons and weather on sleep patterns measured through longitudinal multimodal sensing. npj Digit Med. 2021 Apr 28;4(1):1–15.

63. Cedeño Laurent JG, Williams A, Oulhote Y, Zanobetti A, Allen JG, Spengler JD. Reduced cognitive function during a heat wave among residents of non-air-conditioned buildings: An observational study of young adults in the summer of 2016. PLoS Med. 2018 Jul 10;15(7):e1002605.

64. Li W, Bertisch SM, Mostofsky E, Vgontzas A, Mittleman MA. Associations of daily weather and ambient air pollution with objectively assessed sleep duration and fragmentation: a prospective cohort study. Sleep Medicine. 2020 Nov;75:181–7.

65. Wang C, Liu K, Wang H. The effects of temperature on sleep experience: evidence from China. Applied Economics. 2022 Oct 11;0(0):1–17.

66. van Loenhout JAF, le Grand A, Duijm F, Greven F, Vink NM, Hoek G, et al. The effect of high indoor temperatures on self-perceived health of elderly persons. Environmental Research. 2016;146:27–34.

67. Pandey J, Grandner M, Crittenden C, Smith MT, Perlis ML. Meteorologic factors and subjective sleep continuity: A preliminary evaluation. International Journal of Biometeorology. 2005;49(3):152–5.

68. An R, Yu H. Impact of ambient fine particulate matter air pollution on health behaviors: a longitudinal study of university students in Beijing, China. Public Health. 2018 Jun;159:107–15.

69. Lappharat S, Taneepanichskul N, Reutrakul S, Chirakalwasan N. Effects of bedroom environmental conditions on the severity of obstructive sleep apnea. Journal of Clinical Sleep Medicine. 2018;14(4):565–73.

70. Quinn A, Shaman J. Health symptoms in relation to temperature, humidity, and self-reported perceptions of climate in New York City residential environments. International Journal of Biometeorology. 2017;61(7):1209–20.

71. Yan Y, Lan L, Zhang H, Sun Y, Fan X, Wyon DP, et al. Association of bedroom environment with the sleep quality of elderly subjects in summer: A field measurement in Shanghai, China. Building and Environment. 2022 Jan 15;208:108572.

72. Williams AA, Spengler JD, Catalano P, Allen JG, Cedeno-Laurent JG. Building Vulnerability in a Changing Climate: Indoor Temperature Exposures and Health Outcomes in Older Adults Living in Public Housing during an Extreme Heat Event in Cambridge, MA. International Journal of Environmental Research and Public Health. 2019 Jan;16(13):2373.

73. Xiong J, Lan L, Lian Z, De dear R. Associations of bedroom temperature and ventilation with sleep quality. Science and Technology for the Built Environment. 2020 Oct 20;26(9):1274–84.

74. Xu X, Lian Z, Shen J, Lan L, Sun Y. Environmental factors affecting sleep quality in summer: a field study in Shanghai, China. Journal of Thermal Biology. 2021 Jul 1;99:102977.

75. Milando CW, Black-Ingersoll F, Heidari L, López-Hernández I, de Lange J, Negassa A, et al. Mixed methods assessment of personal heat exposure, sleep, physical activity, and heat adaptation strategies among urban residents in the Boston area, MA. BMC Public Health. 2022 Dec 10;22(1):2314.

76. Zanobetti A, Redline S, Schwartz J, Rosen D, Patel S, O’Connor GT, et al. Associations of PM10 with sleep and sleep-disordered breathing in adults from seven U.S. urban areas. American journal of respiratory and critical care medicine. 2010 Sep;182(6):819– 25.

77. Cassol CM, Martinez D, Da Silva FABS, Fischer MK, Lenz MDCS, Bós ÂJG. Is sleep apnea a winter disease? Meteorologic and sleep laboratory evidence collected over 1 decade. Chest. 2012;142(6):1499–507.

78. Liu WT, Wang YH, Chang LT, Wu CD, Wu D, Tsai CY, et al. The impacts of ambient relative humidity and temperature on supine position-related obstructive sleep apnea in adults. Environmental Science and Pollution Research. 2022 Jul;29(33):50755–50764.

79. Quante M, Wang R, Weng J, Kaplan ER, Rueschman M, Taveras EM, et al. Seasonal and weather variation of sleep and physical activity in 12-14-year-old children. Behav Sleep Med. 2019 Aug;17(4):398–410.

80. Cepeda M, Koolhaas CM, Rooij FJA van, Tiemeier H, Guxens M, Franco OH, et al. Seasonality of physical activity, sedentary behavior, and sleep in a middle-aged and elderly population: The Rotterdam study. Maturitas. 2018 Apr 1;110:41–50.

81. Okamoto-Mizuno K, Tsuzuki K. Effects of season on sleep and skin temperature in the elderly. International Journal of Biometeorology. 2010 Jul;54(4):401–9.

82. Weinreich G, Wessendorf TE, Pundt N, Weinmayr G, Hennig F, Moebus S, et al. Association of short-term ozone and temperature with sleep disordered breathing. European Respiratory Journal. 2015;46(5):1361–9.

83. Ohnaka T, Tochihara Y, Kanda K. Body movements of the elderly during sleep and thermal conditions in bedrooms in summer. Appl Human Sci. 1995 Mar;14(2):89–93.

84. Okamoto-Mizuno K, Tsuzuki K, Mizuno K. Effects of humid heat exposure in later sleep segments on sleep stages and body temperature in humans. Int J Biometeorol. 2005 Mar;49(4):232–7.

85. Obradovich N, Minor K. Identifying and Preparing for the Mental Health Burden of Climate Change. JAMA Psychiatry. 2022 Apr 1;79(4):285–6.

86. Vergunst F, Berry HL, Minor K, Chadi N. Climate Change and Substance-Use Behaviors: A Risk-Pathways Framework. Perspect Psychol Sci. 2022 Nov 28;17456916221132740.

87. Koch M, Matzke I, Huhn S, Gunga HC, Maggioni MA, Munga S, et al. Wearables for Measuring Health Effects of Climate Change-Induced Weather Extremes: Scoping Review. JMIR Mhealth Uhealth. 2022 Sep 9;10(9):e39532.

88. Altena E, Baglioni C, Sanz-Arigita E, Cajochen C, Riemann D. How to deal with sleep problems during heatwaves: practical recommendations from the European Insomnia Network. J Sleep Res. 2022 Sep 8;e13704.

89. Smith CJ. Pediatric Thermoregulation: Considerations in the Face of Global Climate Change. Nutrients. 2019 Sep;11(9):2010.

90. Martínez-Solanas È, López-Ruiz M, Wellenius GA, Gasparrini A, Sunyer J, Benavides FG, et al. Evaluation of the Impact of Ambient Temperatures on Occupational Injuries in Spain. Environ Health Perspect. 2018 Jun;126(6):067002.

91. Burke M, González F, Baylis P, Heft-Neal S, Baysan C, Basu S, et al. Higher temperatures increase suicide rates in the United States and Mexico. Nature Climate Change. 2018 Aug;8(8):723–9.

92. Moulton BR. Random group effects and the precision of regression estimates. Journal of Econometrics. 1986 Aug 1;32(3):385–97.

93. Berrang-Ford L, Sietsma AJ, Callaghan M, Minx JC, Scheelbeek PFD, Haddaway NR, et al. Systematic mapping of global research on climate and health: a machine learning review. The Lancet Planetary Health. 2021 Aug 1;5(8):e514–25.

94. Irish LA, Kline CE, Gunn HE, Buysse DJ, Hall MH. The Role of Sleep Hygiene in Promoting Public Health: A Review of Empirical Evidence. Sleep Med Rev. 2015 Aug;22:23–36.

95. Ravanelli NM, Jay O. Electric fan use in heat waves: Turn on or turn off? Temperature. 2016 Jul 2;3(3):358–60.

96. Gagnon D, Crandall CG. Electric fan use during heat waves: Turn off for the elderly? Temperature. 2017 Apr 3;4(2):104–6.

97. Brown HA, Topham TH, Clark B, Smallcombe JW, Flouris AD, Ioannou LG, et al. Seasonal Heat Acclimatisation in Healthy Adults: A Systematic Review. Sports Med. 2022 Sep;52(9):2111–28.

98. Morrison SA. Moving in a hotter world: Maintaining adequate childhood fitness as a climate change countermeasure. Temperature. 2022 Aug 4;0(0):1–19.

99. Eggeling J, Rydenfält C, Kingma B, Toftum J, Gao C. The usability of ClimApp: A personalized thermal stress warning tool. Climate Services. 2022 Aug 1;27:100310.

100. Chevance G, Perski O, Hekler EB. Innovative methods for observing and changing complex health behaviors: four propositions. Transl Behav Med. 2021 Mar 16;11(2):676–685.

101. Gaston SA, Singh R, Jackson CL. The Need to Study the Role of Sleep in Climate Change Adaptation, Mitigation, and Resiliency Strategies across the Life Course. Sleep. 2023 Mar 13;zsad070.

102. Iungman T, Cirach M, Marando F, Barboza EP, Khomenko S, Masselot P, et al. Cooling cities through urban green infrastructure: a health impact assessment of European cities. The Lancet. 2023 Feb 18;401(10376):577–89.

103. Bowler DE, Buyung-Ali L, Knight TM, Pullin AS. Urban greening to cool towns and cities: A systematic review of the empirical evidence. Landscape and Urban Planning. 2010 Sep 15;97(3):147–55.

104. Donat MG, Alexander LV, Yang H, Durre I, Vose R, Dunn RJH, et al. Updated analyses of temperature and precipitation extreme indices since the beginning of the twentieth century: The HadEX2 dataset. Journal of Geophysical Research: Atmospheres. 2013;118(5):2098–118.

105. Cox DTC, Maclean IMD, Gardner AS, Gaston KJ. Global variation in diurnal asymmetry in temperature, cloud cover, specific humidity and precipitation and its association with leaf area index. Global Change Biology. 2020;26(12):7099–111.

106. Wang J, Chen Y, Tett SFB, Yan Z, Zhai P, Feng J, et al. Anthropogenically-driven increases in the risks of summertime compound hot extremes. Nat Commun. 2020 Feb 11;11(1):528.

107. Huang K, Li X, Liu X and Seto KC. Projecting global urban land expansion and heat island intensification through 2050. Environ Res Lett. 2019;14 114037.

