## Supplementary material for "A systematic review of ambient heat and sleep in a warming climate": supplemetary file

PUBMED
((( "Sleep/economics"[Mesh] OR "Sleep/epidemiology"[Mesh] OR "Sleep/methods"[Mesh] OR "Sleep/mortality"[Mesh] OR "Sleep/nursing"[Mesh] OR "Sleep/organization and administration"[Mesh] OR "Sleep/pathology"[Mesh] OR "Sleep/pharmacology"[Mesh] OR "Sleep/physiology"[Mesh] OR "Sleep/prevention and control"[Mesh] OR "Sleep/psychology"[Mesh] OR "Sleep/rehabilitation"[Mesh] OR "Sleep/statistics and numerical data"[Mesh] )) OR ( "Sleep Initiation and Maintenance Disorders/epidemiology"[Mesh] OR "Sleep Initiation and Maintenance Disorders/mortality"[Mesh] OR "Sleep Initiation and Maintenance Disorders/nursing"[Mesh] OR "Sleep Initiation and Maintenance Disorders/prevention and control"[Mesh] OR "Sleep Initiation and Maintenance Disorders/psychology"[Mesh] OR "Sleep Initiation and Maintenance Disorders/rehabilitation"[Mesh] OR "Sleep Initiation and Maintenance Disorders/therapy"[Mesh] )) AND ( "Weather"[Mesh] OR "Extreme Cold Weather"[Mesh] OR "Extreme Hot Weather"[Mesh] OR "Extreme Weather"[Mesh] OR "Temperature/physiology"[Mesh] OR "Temperature/prevention and control"[Mesh] OR "Temperature/psychology"[Mesh])

SCOPUS

TITLE-ABS-KEY ( ( *weather* OR *"temperature"* OR *"heat index"* OR *"wet bulb globe"* AND ( *sleep* OR *insomnia* ) AND NOT ( *mice* OR *animal* OR *murin* OR *"body temperature"* OR *"skin temperature"* ) ) ) AND ( LIMIT-TO ( SRCTYPE , *"j"* ) ) AND ( LIMIT-TO ( DOCTYPE , *"ar"* ) ) AND ( LIMIT-TO ( SUBJAREA , *"MEDI"* ) OR LIMIT-TO ( SUBJAREA , *"PSYC"* ) OR LIMIT-TO ( SUBJAREA , *"COMP"* ) OR LIMIT-TO ( SUBJAREA , *"ENVI"* ) ) AND ( LIMIT-TO ( LANGUAGE , *"English"* ) ) AND ( EXCLUDE ( SUBJAREA , *"BIOC"* ) OR EXCLUDE ( SUBJAREA , *"ENGI"* ) )

JSTOR
(ab:(sleep) OR ab:(insomnia)) AND ab:(temperature)

PsyInfo, greenfile, eric, georef
(AB sleep OR AB ( insomnia or sleep disorders or sleep disturbance )) AND (AB temperature OR AB weather OR AB heat index OR AB wet bulb OR AB ambient temperature) NOT (AB skin temperature NOT AB body temperature NOT AB ( mouse or mice or rats ) NOT AB murine)
